# neuroGPT-X: Towards an Accountable Expert Opinion Tool for Vestibular Schwannoma

**DOI:** 10.1101/2023.02.25.23286117

**Authors:** Edward Guo, Mehul Gupta, Sarthak Sinha, Karl Rössler, Marcos Tatagiba, Ryojo Akagami, Ossama Al-Mefty, Taku Sugiyama, Philip E. Stieg, Gwynedd E. Pickett, Madeleine de Lotbiniere-Bassett, Rahul Singh, Sanju Lama, Garnette R. Sutherland

## Abstract

**Background:** The global launch of ChatGPT on November 30, 2022 has sparked widespread public interest in large language models (LLMs), and interest in the medical community is growing. Indeed, recent preprints on medRxiv have examined ChatGPT and GPT-3 in the context of standardized exams, such as the United States Medical Licensing Examination. These studies demonstrate modest performance relative to national averages. In this work, we enhance OpenAI’s GPT-3 model through zero-shot learning, anticipating that it outperforms experienced neurosurgeons in written question-answer tasks for common clinical and surgical questions on vestibular schwannoma. We aimed to address LLM accountability by including in-text citations and references to the responses provided by GPT-3.

**Methods:** The analysis involved (i) creating a dataset through web scraping, (ii) developing a chat-based platform called neuroGPT-X, (iii) enlisting expert neurosurgeons across international centers to create and answer questions and evaluate responses, and (iv) analyzing the evaluation results on the management of vestibular schwannoma. The survey had a blinded and unblinded phase. In the blinded phase, a neurosurgeon with 30+ years of experience curated 15 questions regarding common clinical and surgical contexts of vestibular schwannoma. Then, four neurosurgeons, ChatGPT (January 30, 2023 model, aka *naive* GPT), and a context-enriched GPT model independently provided their responses. Three experienced neurosurgeons blindly evaluated the responses for accuracy, coherence, relevance, thoroughness, speed, and overall rating. Then, all seven neurosurgeons were unblinded to all responses and provided their thoughts on the potential of expert LLMs in the clinical setting.

**Findings:** Both the naive and content-enriched GPT models provided faster responses to the standardized question set (p<0.01) than expert neurosurgeon respondents. Moreover, responses from both models were consistently non-inferior in accuracy, coherence, relevance, thoroughness, and overall performance, and were often rated higher than expert responses. Importantly, context enrichment of GPT with relevant scientific literature did not significantly affect speed (p>0.999) or performance across the aforementioned domains (p>0.999). Of interest, all expert surgeons expressed concerns about the reliability of GPT in accurately addressing the nuances and controversies surrounding the management of vestibular schwannoma. Further, we developed neuroGPT-X, a chat-based platform designed to provide point-of-care clinical support and mitigate limitations of human memory. neuroGPT-X incorporates features such as in-text citations and references to enable accurate, relevant, and reliable information in real-time.

**Interpretation:** A context-enriched GPT model provided non-inferior responses compared to experienced neurosurgeons in generating written responses to a complex neurosurgical problem for which evidence-based consensus for management is lacking. We show that context enrichment of LLMs is well-suited to transform clinical practice by providing subspecialty-level answers to clinical questions in an accountable manner.

**Research in Context:** *Evidence before this study:* We searched PubMed for “(vestibular schwannoma OR acoustic schwannoma) AND (GPT-3 OR Generative Pretrained Transformer OR large language model)” with no filters and identified no relevant articles. We then searched PubMed using the string “(subspecialty OR neurosurgery OR physician) AND (GPT-3 OR Generative Pretrained Transformer OR large language model) AND (fine-tuning OR context enrichment)” with no filters and identified three studies. One study noted that domain-specific knowledge enhanced pre-trained language models.

*Added value of this study:* To our knowledge, this is the first study to show the non-inferiority of a context-enriched LLM in a question-answer task on common clinical and surgical questions compared to experienced neurosurgeons worldwide, determined by their neurosurgical colleagues. Furthermore, we developed the first online platform incorporating an LLM, chat memory, in-text citations, and references regarding comprehensive vestibular schwannoma management. To assess the model’s performance, a neurosurgeon with 30+ years of experience managing patients with vestibular schwannoma curated 15 questions to the model, ChatGPT, and four international expert neurosurgeons. A separate, blinded group of three expert neurosurgeons assessed these answers for accuracy, coherence, relevance, thoroughness, speed, and overall rating. This study demonstrated the capability of context-enriched LLMs as point-of-care informational aids. Importantly, all expert surgeons raised questions regarding the nuances and role of human experience and intuition that GPT may not capture in generating opinions or recommendations.

*Implications of all the available evidence:* The present study, with its subspecialist-level performance and interpretable results, suggests that context-enriched LLMs show promise as a point-of-care medical resource. Evaluations from experienced neurosurgeons showed that a context-enriched GPT model was rated similarly to neurosurgeon responses across evaluation domains in this study. This work serves as a springboard for expanding this tool into more medical specialties, incorporating evidence-based clinical information, and developing expert-level dialogue surrounding LLMs in healthcare.

## Introduction

Over the past several decades, information storage and retrieval have shifted from textbooks and journals to digital resources. Web-based knowledge, including mobile applications, has provided the medical community rapid access to an ever-increasing body of knowledge.^1^ While physicians may access information and guidelines using search engines, results are usually high-volume, necessitating selective data extraction into clinically relevant information. In addition, physicians are continually required to remain up-to-date on their knowledge and skills, relying on experience and the latest published standards of care.^2^ With more than one million publications a year (two papers/minute) added to online databases in the biomedical field alone, the current mode of learning, teaching, and practicing medicine is rapidly becoming volume-intensive and memory-prohibitive.^1,3,4^ Such an approach to education is time-consuming, with information often segregated, variable, or incomplete, requiring further in-depth review and analysis to achieve optimal results.^4^ In this era of rapid medical advancement, it is imperative that physicians have access to the latest and most relevant information to ensure the best outcomes for their patients.

Large language models (LLMs), such as Generative Pretrained Transformer (GPT),^5^ show promise as clinical reasoning tools. LLMs are trained on massive amounts of text data to generate human-like language and perform tasks such as text generation, question answering, and language translation.^5^ The release of ChatGPT in November 2022, which features a chat-like interface and human-like conversation skills, highlights the potential of LLMs in fields that require complex decision-making. This is exemplified by recent studies that demonstrate the performance of ChatGPT and GPT on standardized exams, such as the United States Medical Licensing Examination^6^ and the National Conference of Bar Examiners Bar Exam.^7^

The use of LLMs as a point-of-care medical resource for subspecialty physicians has yet to be investigated. Despite its potential, issues such as interpretability and accountability have limited the application of LLMs in high-risk environments. However, context enrichment and zero-shot learning have shown marked domain-specific performance improvements,^8^ making this an area of considerable promise. Combining zero-shot learning and publication references with LLM responses may address interpretability and accountability concerns, thereby improving the clinical utility of these models.

To establish whether LLMs can assist in educating residents and delivering treatment options at an experienced physician level, it would be important to challenge LLMs with a complex and controversial disease paradigm. One such example is acoustic/vestibular schwannoma, which bears nuances of intricate anatomical landmarks, diverse diagnostic modalities, global consensus treatment approaches, and surgeon expertise, whereby treatment and outcomes may vary between centers of excellence.^9^ Staying abreast of literature and guidelines while maintaining technical excellence may be challenging, requiring a balance between knowledge extraction and reasoning.

In this study, we make three key contributions toward assessing LLMs as a clinical tool. Firstly, we develop a framework to enrich GPT with context relevant to vestibular schwannoma. Secondly, we compare the performance of ChatGPT, termed *naive* GPT, and a context-enriched GPT model against leading neurosurgical experts worldwide. Finally, we introduce a proof-of-concept clinical tool, *neuroGPT-X*, which incorporates working memory and sources with its responses in a web-based chat platform, aimed at addressing the challenges of LLMs in a clinical setting (e.g., interpretability, reliability, accountability, and safety). Here we hypothesize that a well-trained, context-enriched GPT will perform similar to or better than expert surgeons in answering questions commonly encountered in day-to-day practice.

## Methods

### Dataset Curation

Terminologies unique to vestibular schwannoma were used to obtain articles and abstracts via web scraping from Wikipedia and PubMed. The Wikipedia Python application programming interface (API) was used to scape all webpages from the starting webpage of “Vestibular Schwannoma,” and the content from each page was extracted by heading. The findpapers Python application^10^ was used to scrape abstracts from journals and conference proceedings related to vestibular schwannoma into a structured JSON format from PubMed from January 1, 2000, until January 1, 2023. The query was *([vestibular schwannoma] OR [acoustic neurilemmoma] OR [perineural fibroblastoma] OR [neurinoma of the acoustic nerve] OR [neurofibroma of the acoustic nerve] OR [schwannoma of the acoustic nerve])*. All journals flagged as potentially predatory in the metadata of the findpapers application output were removed based on Beall’s List of potentially predatory journals and publishers.

A structured dataset was compiled with these column names: Title, Heading, Content, Authors, Tokens, and Embeddings. For Wikipedia articles, “Title” was the name of the article, “Heading” was the article title, and “Authors” was set as “Wikipedia.” For PubMed abstracts, “Title” was “Vestibular Schwannoma,” “Heading” was the publication title, and “Authors” were the contributing author(s). The “Content” column included the scraped Wikipedia section or PubMed abstract. The “Tokens” column had the number of tokens for each row in the “Content” column computed using the GPT2TokenizerFast function provided by OpenAI. These tokens were used to calculate the maximum input length to provide to the OpenAI GPT API. The “Embeddings” column included embeddings from the “Content” column computed using the OpenAI embedding model, text-embedding-ada-002. These embeddings were used for thematic analysis and search purposes to find the most similar documents to a user’s input, providing context enrichment for the OpenAI GPT-3 API in a zero-shot learning fashion. We call this model context-enriched GPT.

### Thematic Analysis

Thematic analysis was performed using the dataset embeddings. K-means clustering classified clusters of thematic similarity; the clusters were chosen based on the elbow method and silhouette score for clusters ranging from 2-25. The t-SNE algorithm was employed for data visualization with two components, a perplexity of the square root of the number of rows in the structured dataset, PCA as an initializer, and a learning rate of 200. The InstructGPT Davinci model,^11^ text-davinci-003, was used to perform thematic analysis on a randomly sampled subset of articles from each cluster. Each cluster’s theme was summarized into keywords using the same text-davinci-003 model with hyperparameters of temperature = 0, max_tokens = 64, top_p = 1, frequency_penalty = 0, and presence_penalty = 0. The prompt for the thematic analysis was *“What do the following abstracts have in common?”* and the prompt for the summarization was *“Summarize this phrase into keywords*.*”*

### Context Enrichment with Prompt Construction

User input was enriched with a priming prompt and content for all requests to the OpenAI GPT API. Standard prompts were constructed using the following structure:

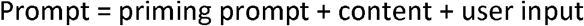

The priming prompt was, *“Pretend you are a physician writing an exam. Please answer every question to the highest degree of medical accuracy. Provide detailed reasoning for your answer. Use the context provided to supplement your knowledge base*.*”* The content comprised Wikipedia article sections and PubMed abstracts sorted by cosine similarity had a maximum token length of 2000. Context-enriched GPT output had a maximum token length of 2000.

Cosine similarity between user input and structured dataset content was used to obtain relevant context enrichment for GPT via OpenAI’s embedding model, text-embedding-ada-002. More precisely, given user input as an embedding vector ***U*** and an embedding within a structured dataset ***E***, the cosine similarity between ***U*** and ***E*** was defined as the dot product between ***U*** and ***E*** divided by the product of the magnitudes of ***U*** and ***E***.

### Web Application, Memory, and Inclusion of Sources

A web application was developed to create a user chat interface for the context-enriched GPT model using the Python Flask microweb framework^12^ paired with HTML, CSS, and Javascript. We call this platform *neuroGPT-X*. User messages are permitted up to 1500 characters (∼210-380 words). Key features of neuroGPT-X include conversation memory of the current session, in-text citations and references for its responses, restriction of conversations to relevant medical topics, timestamps for all messages, animated loading icons while neuroGPT-X computes its response, and prevention of user input spam.

Memory was implemented using Conversation Summary Buffer Memory from LangChain, a framework that provides a standard memory interface with LLMs. This type of memory summarizes both new and old interactions in memory and keeps a buffer of recent interactions. In our implementation, the buffer memory had a maximum token limit of 750. Logistically, we save a serialized Conversation Summary Buffer Memory via the Python pickle module after neuroGPT-X completes its response, and we load the memory upon new user inputs. This memory file is deleted after the user completes or terminates each session.

On the backend of neuroGPT-X, the prompt was constructed with the primer:

*“Pretend you are a specialist physician. Answer every question to the highest degree of medical accuracy. Provide detailed reasoning for your answer. Use the context provided to supplement your knowledge base. Cite the relevant context in IEEE style using the enumerated context before punctuation, e*.*g*., *[1], [3, 5]. Ignore the context if it is not relevant to the question. Do not create fake citations or more citations than were provided. Write ‘This is outside the scope of my functionality*.*’ if the question does not relate to vestibular schwannoma or neurosurgery*.*”*

The context was provided using an enumerated list of relevant articles sorted by cosine similarity to the user input up to a token length of 1500. The neuroGPT-X model was given API completion parameters of temperature = 0, model = text-davinci-003, max_tokens = 1500, and top_p = 1.

### Study Methodology

A neurosurgeon with 30+ years of experience with active clinical practice in vestibular schwannoma curated 15 general questions involving anatomy, surgical management, imaging, clinical contexts, and genetic predispositions (Supplementary Table 1). These questions were answered by the (i) January 30, 2023, ChatGPT model, termed naive GPT, (ii) context-enriched GPT, and (iii) four experienced neurosurgeons active in the treatment of vestibular schwannoma, also called *experts* here on. The context-enriched GPT model had API completion parameters of temperature = 0, model = text-davinci-003, max_tokens = 2000, and top_p = 1.

**Table 1.**
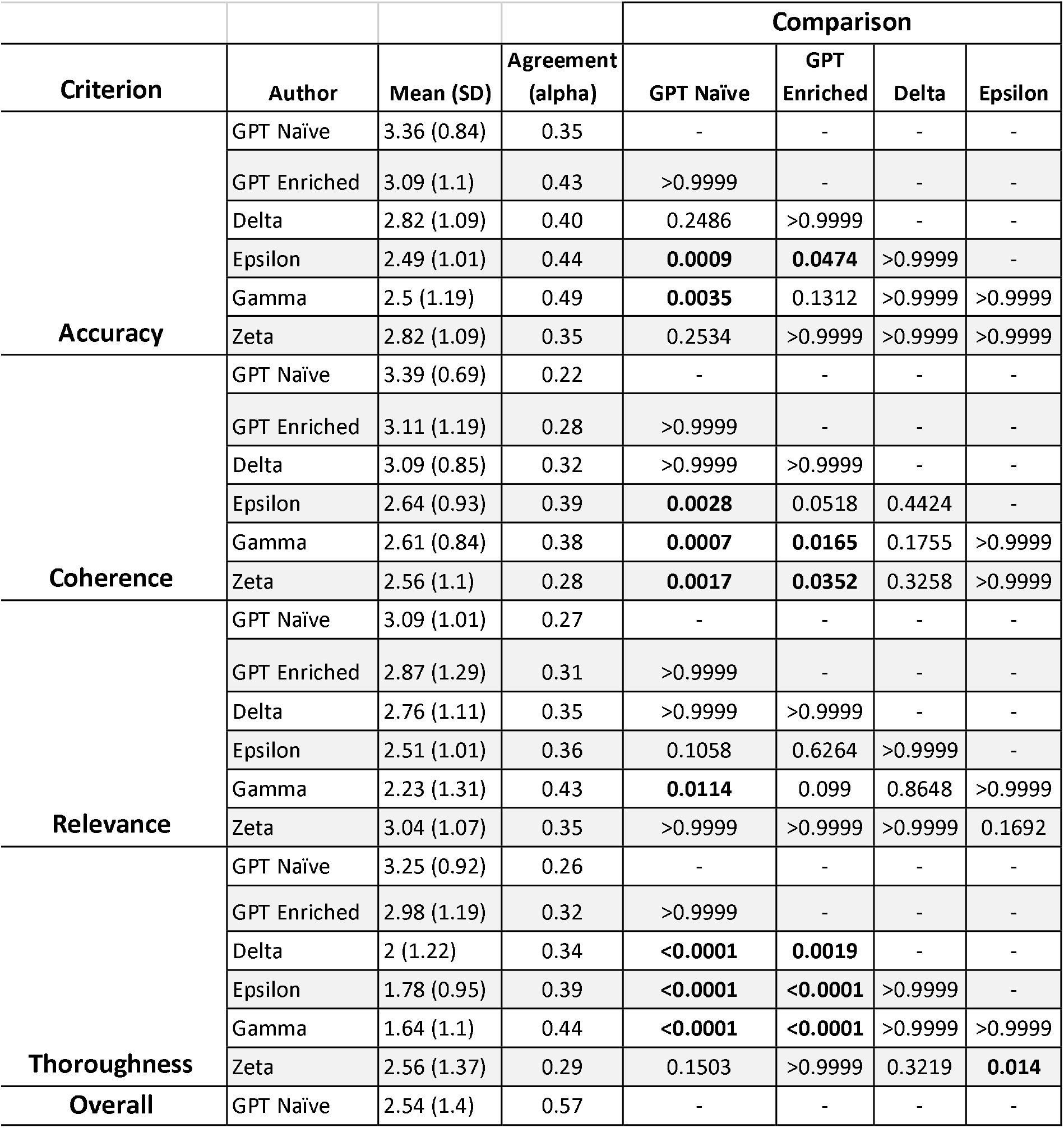
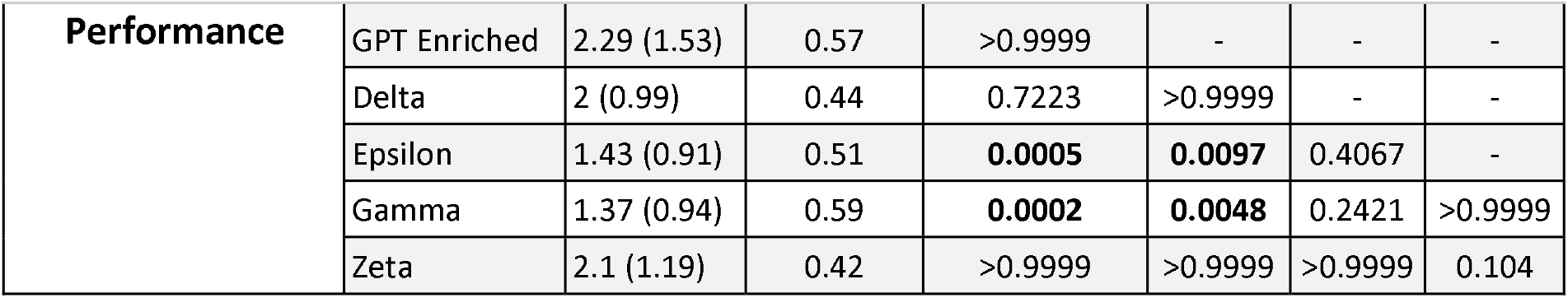
Blinded evaluation scores from three expert neurosurgeons. Krippendorff’s alpha was used to quantify interrater reliability. Kruskal-Wallis tests were used to compare pairwise differences, with multiple comparison-adjusted p-values shown. Responses to 15 curated questions were measured by accuracy, coherence, thoroughness, relevance, and overall score. Gamma, Delta, Epsilon, and Zeta are the neurosurgeons.

The four expert surgeons were asked to answer the 15 curated questions ad hoc as if speaking to residents or patients while timing themselves. The same questions were independently posed to naive GPT and context-enriched GPT, and response times were recorded. The experts, naive GPT, and context-enriched GPT responses were reviewed and compiled by an independent investigator. The evaluation of these answers was then divided into a *blinded* and *unblinded* phase.

In the *blinded* phase, three independent neurosurgeons blindly evaluated the responses from naive GPT, context-enriched GPT, and the four experts. The evaluation metrics included accuracy, coherence, relevance, thoroughness, and overall rating on a 0-4 Likert scale, with 4 indicating a better answer (**Supplementary Table 2**). The evaluators were also asked whether they thought the response was provided by an expert or naive GPT/context-enriched GPT.

**Table 2.**
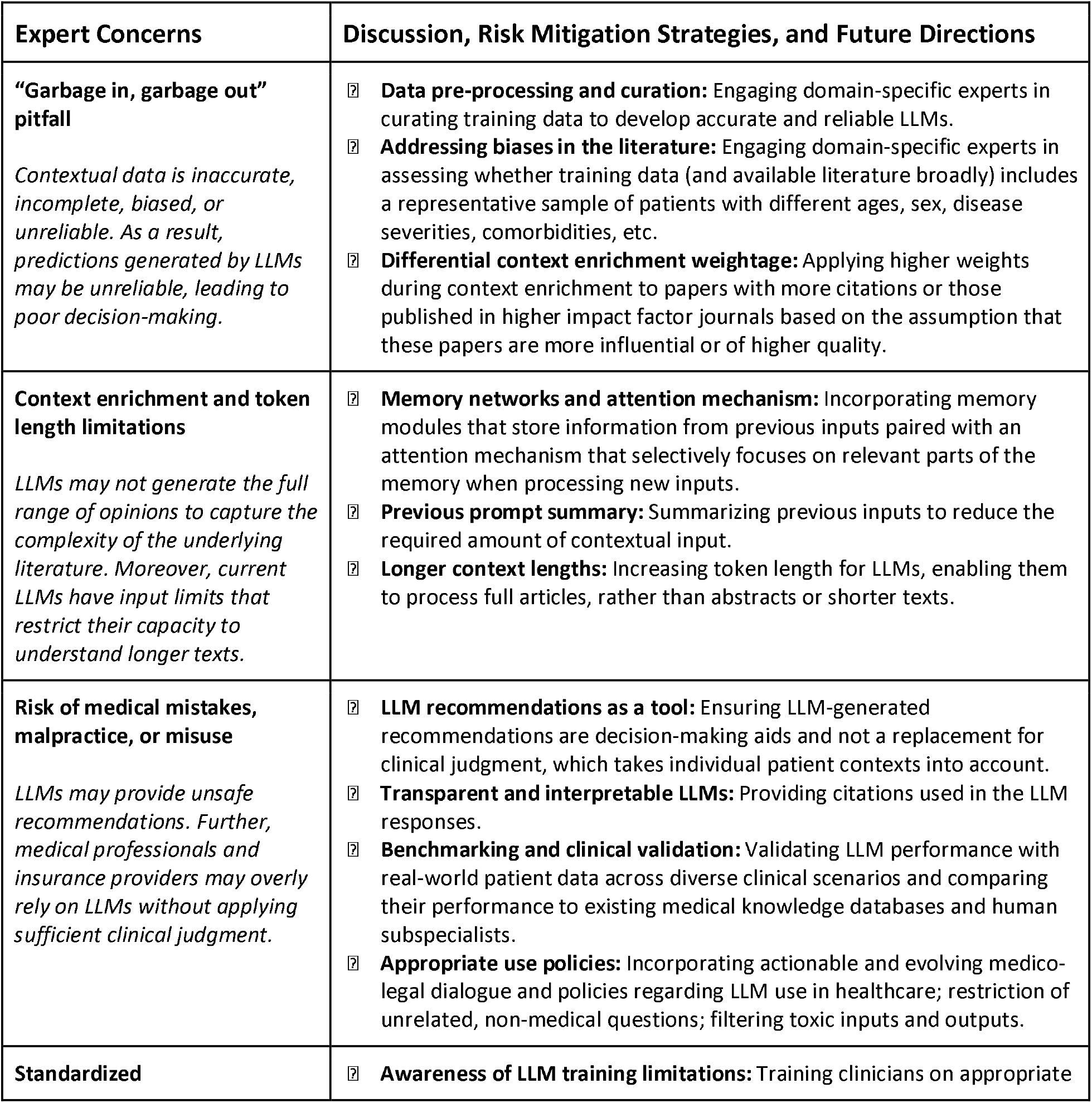
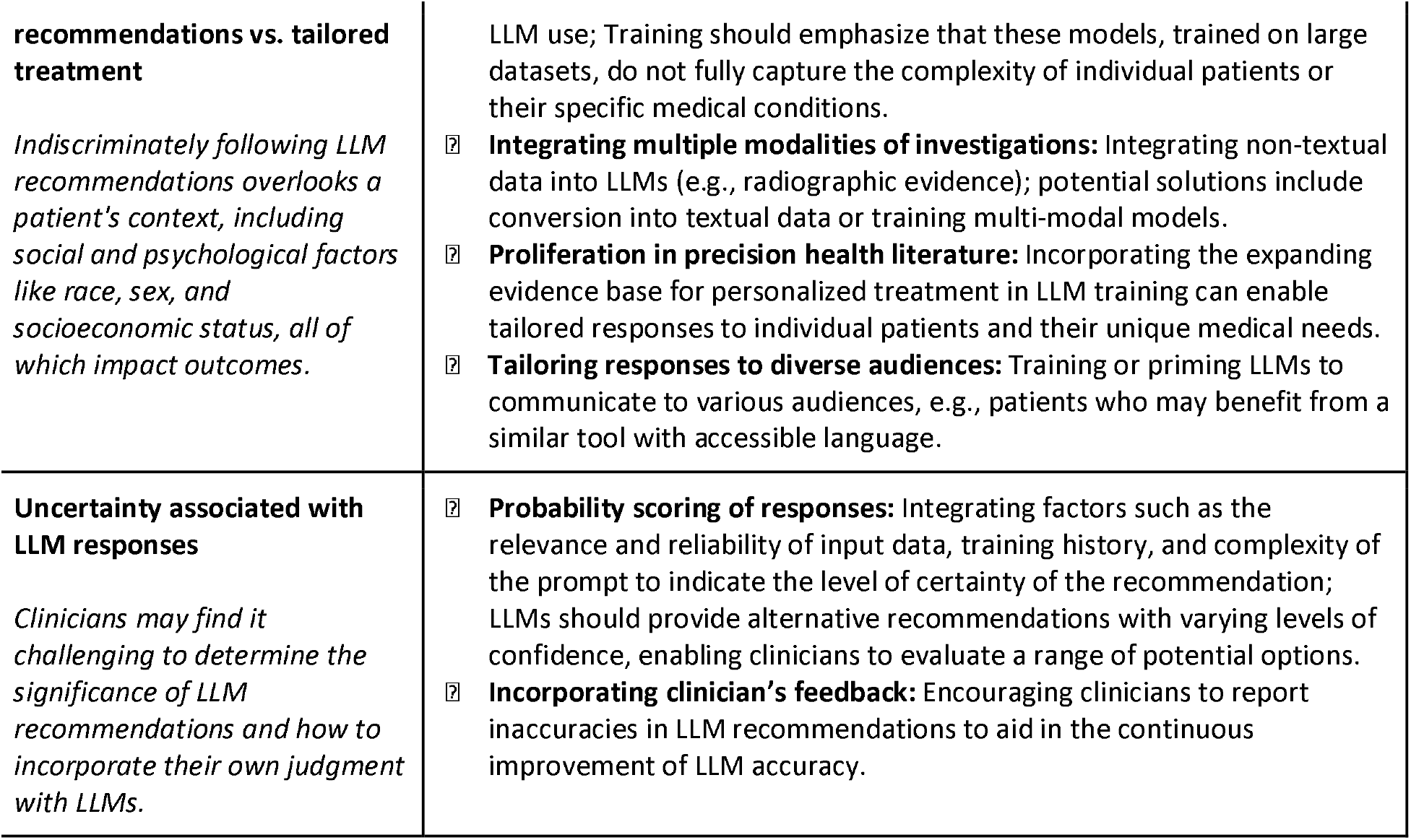
Expert concerns from the unblinded phase of this study. The authors discussed these concerns, and we present a summary of consensus recommendations.

After initial evaluations, the experts and evaluators were *unblinded* to naive and context-enriched GPT responses. In this phase, all seven expert surgeons were asked questions regarding GPT-generated responses, including overall satisfaction, the likelihood of use in the clinic, value, and the likelihood they would recommend the tool to colleagues via a Likert scale from 0-4, with 4 indicating a better answer (**Supplementary Table 2**). The evaluators were also asked long-form questions about their thoughts on GPT in clinical practice.

### Statistical Analysis

Statistical analyses were performed using Python 3.9 and visualized in GraphPad Prism. Statistical significance was set at an alpha of 0.05. Krippendorff’s alpha was computed to measure interrater agreement between evaluators in the blinded phase. In the blinded phase, aggregate scores and timing measures were compiled across all questions for naive GPT, context-enriched GPT, and neurosurgeon experts. The normality of the data was evaluated using the Shapiro-Wilk test, and the Kruskal-Wallis test, followed by adjustments for multiple comparisons if significant, was used to compute statistical differences between the responses. Data was visualized corresponding to the dimension of evaluation metrics, i.e., accuracy, coherence, relevance, thoroughness, and overall rating. In the affective phase, piecharts were used to visualize the expert perception of the clinical utility of GPT-generated responses.

## Results

### Dataset Curation, Thematic Analysis, and neuroGPT-X

Web scraping using the Wikipedia API returned 157 articles, further divided into 1,659 sections separated by headings. The findpapers Python application returned 3,093 publications from PubMed, 12 publications from ACM Digital Library, 6 publications from arXiv, 2 publications from bioRxiv, and 1 paper from medRxiv. Four publications flagged as published in potentially predatory journals were removed from the PubMed search. Thus, the total number of Wikipedia articles and publications was 3,267. **Fig. 1** visualizes these results.

**Figure 1.**
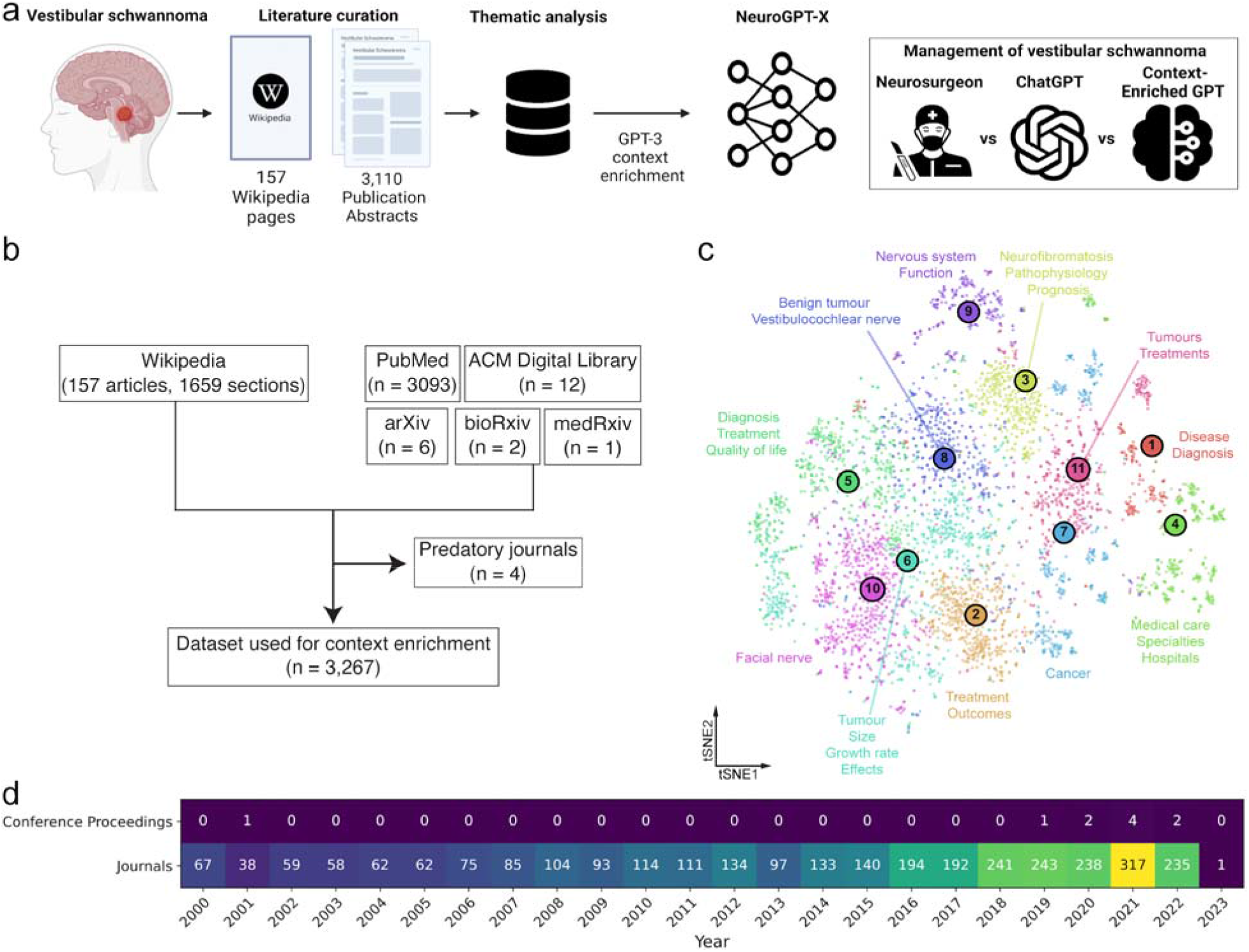
Overview of the study pipeline and dataset characteristics. (a) The data processing pipeline and the three comparisons in this study. (b) Data inclusion and exclusion criteria. (c) Thematic analysis of the embedding vectors of the vestibular schwannoma dataset. Clusters were computed using K-means clustering. The InstructGPT Davinci model was used to classify each cluster. **Supplementary Table 2** provides detailed thematic analysis results. (d) Peer-reviewed abstract publication numbers by year and type.

In thematic analyses, the elbow method returned an optimal number of clusters of 11 with a silhouette coefficient of 859.35. The thematic analysis produced a thematic analysis of each cluster, and the keywords associated with clusters 1-11 are in **Supplementary Table 3. Fig. 2** shows the neuroGPT-X user interface and an example of an interaction with the platform. An entire conversation, including answers to the 15 curated questions in this study, is included in the supplementary data (**Supplementary Table 4**). The website can be accessed here: https://neurogpt-x.azurewebsites.net/.

**Figure 2.**
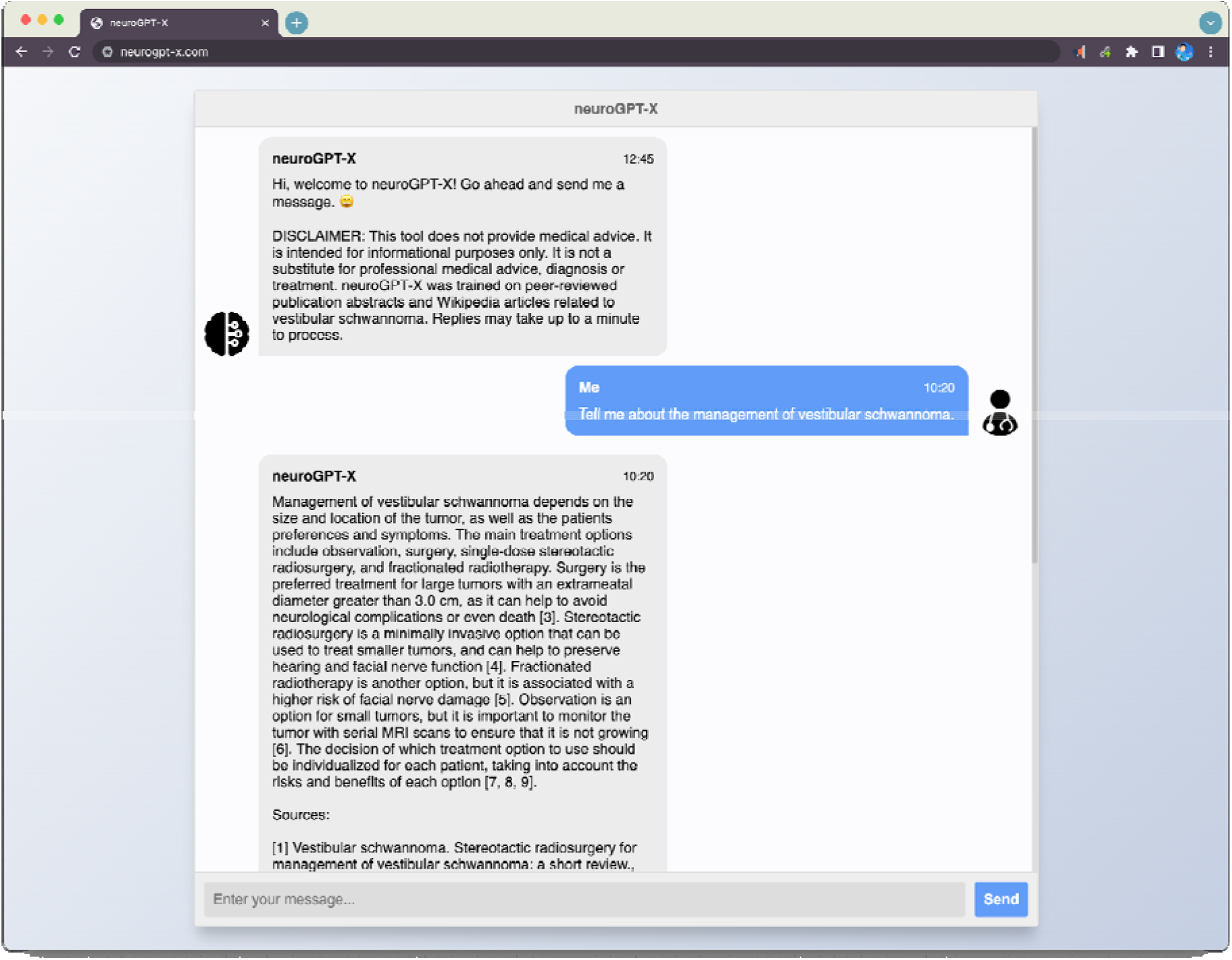
The neuroGPT-X chat interface. Features include conversation memory of the current session, references for the information it provides, restriction of conversations to relevant medical topics, timestamp for each message, animated loading icons while responses are computed, and prevention of user input spam. The website can be accessed here: https://neurogpt-x.azurewebsites.net/.

### Evaluation of Naive GPT, Context-Enriched GPT, and Expert Neurosurgeons

The average response time aggregated across all questions was 63.33 ± 66.08 s for neurosurgeon gamma, 30.00 ± 30.88 s for neurosurgeon delta, 42.00 ± 22.1 s for neurosurgeon epsilon, 134.67 ± 187.15 s for neurosurgeon zeta, 49.03 ± 10.67 s for naive GPT, and 16.67 ± 9.29 s for enriched GPT. The timing normalized by character count was 513.9 ± 140 ms/character for neurosurgeon gamma, 380.7 ± 820 ms/character for neurosurgeon delta, 694.9 ± 710 ms/character for neurosurgeon epsilon, 365.0 ± 240 ms/character for neurosurgeon zeta, 36.5 ± 10 ms/character for naive GPT, and 19.8 ± 10 ms for neuroGPT-X. Pairwise comparisons of normalized response speed demonstrated that both models were significantly faster than the expert neurosurgeon responses (p<0.01), with both models having very similar response times (p>0.999). These results are visualized in **Fig. 3A** and tabulated in **Supplementary Table 5**.

**Figure 3.**
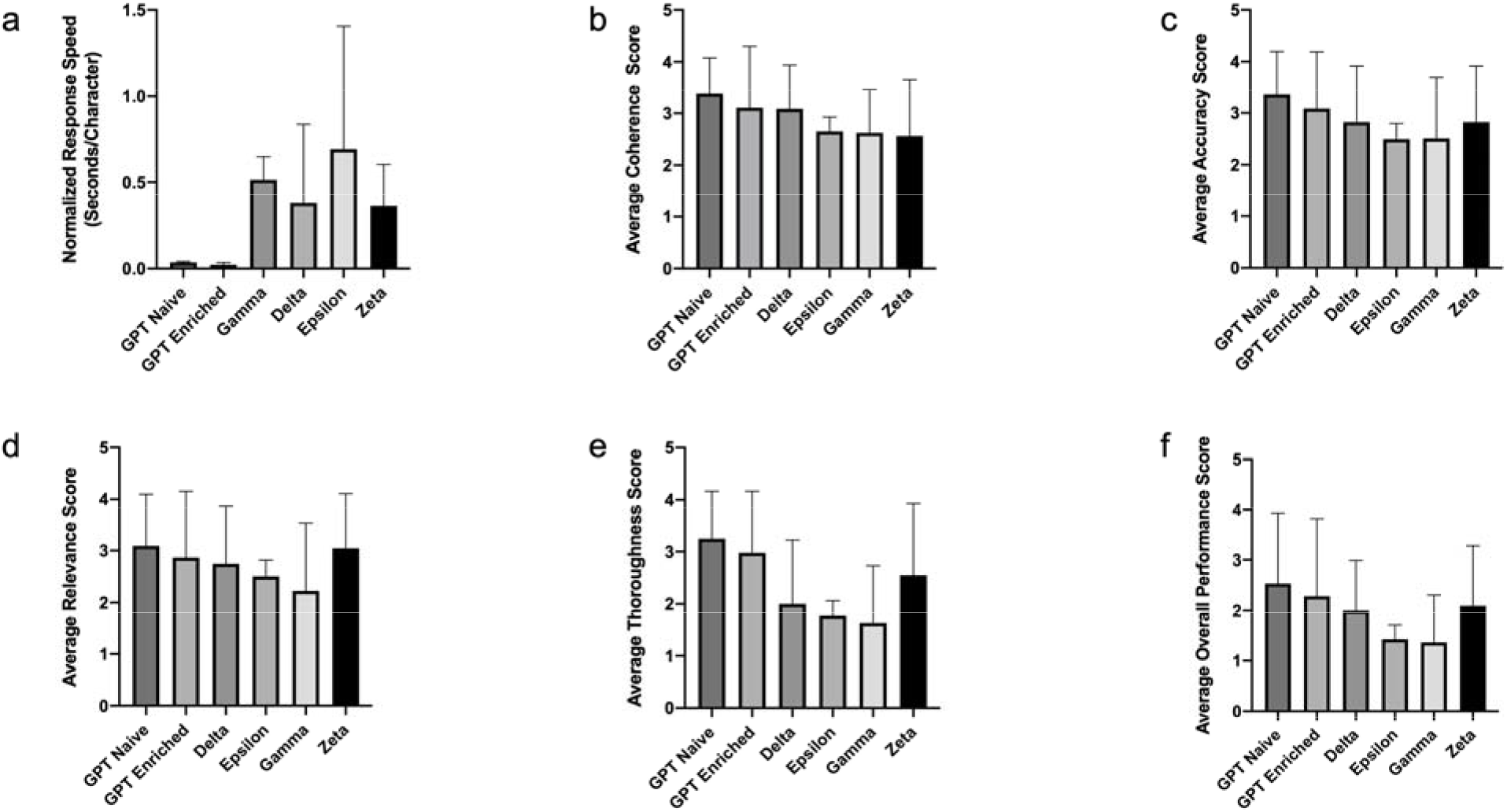
Blinded evaluation scores from three expert neurosurgeons. Responses to 15 curated questions were measured by (a) normalized response speed by response character length, (b) coherence, (c) accuracy, (d) relevance, (e) thoroughness, and (f) overall performance score. Gamma, Delta, Epsilon, and Zeta are the neurosurgeons. Error bars represent standard deviation.

Evaluations by three additional neurosurgeons demonstrated that the naive and enriched GPT model was non-inferior and often superior to the responses obtained from expert neurosurgeons. Kruskal-Wallis one-way ANOVA analysis showed significant differences in performance across accuracy, coherence, relevance, thoroughness, and overall performance metrics (p<0.0001). To assess pairwise performance differences across responses, Kruskal-Wallis tests corrected for multiple comparisons demonstrated consistently non-inferior performance of the naive and enriched GPT models compared to expert neurosurgeons across all metrics. The models outperformed most expert neurosurgeon responses for coherence and thoroughness. Finally, the models had weaker scores in relevance, though both remained non-inferior to expert neurosurgeon responses. A tabular summary of the results can be seen in **Table 1** and is visualized in **Fig. 3B-F**. Krippendorff’s alpha was 0.41 ± 0.05 (±standard deviation) for accuracy, 0.31 ± 0.03 for coherence, 0.35 ± 0.02 for relevance, 0.35 ± 0.05 for thoroughness, and 0.52 ± 0.01 for overall performance (**Table 1**). A visualization of the unblinded affective survey results is shown in **Fig. 4**.

**Figure 4.**
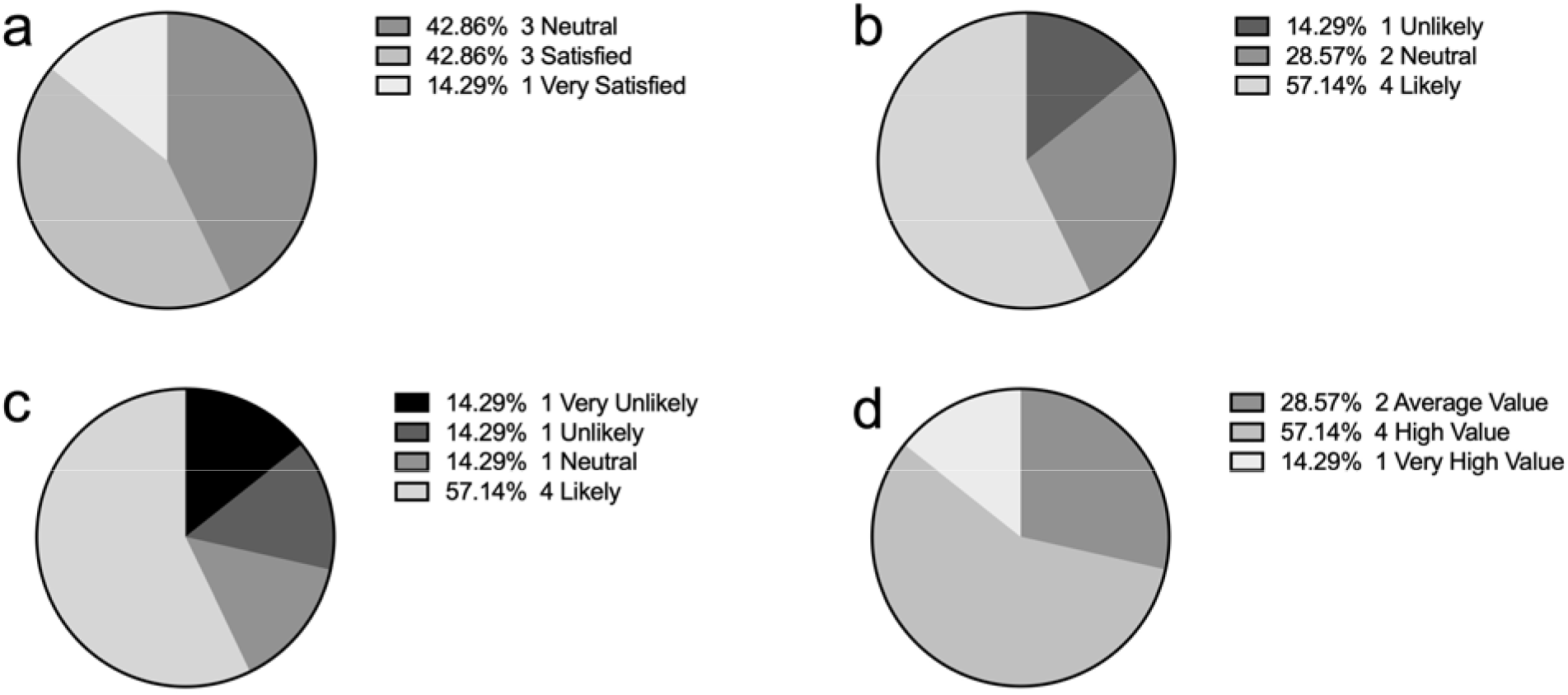
Unblinded survey metrics of all seven neurosurgeon experts and evaluators regarding a context-enriched GPT model measuring (a) overall satisfaction, (b) likelihood of recommendation to colleagues, (c) likelihood of use in the clinic, and (d) perceived value of the tool relative to current resources.

## Discussion

In this study, we demonstrate the non-inferiority of LLM-based model responses to 15 common subspecialist-level questions on vestibular schwannoma compared to experienced neurosurgeons. Moreover, we enriched GPT-3 with relevant, publicly available information via filtered abstracts from scientific publications to supplement the model’s knowledge base and provide reference material. To elicit expert-level responses from the model, a distinct priming prompt was employed, which varied from the prompt provided to the neurosurgeons. To bolster accessibility, functionality, and safety, we developed *neuroGPT-X*, incorporating a zero-shot learning framework with the GPT-3 API into a web application enriched with features. These features included conversation memory, in-text citations, full references, restriction of conversations to relevant medical topics, conversation timestamps, and spam prevention. To our knowledge, this is the first study to show subspecialist-level performance of a context-enriched LLM.

The observation that response coherence and thoroughness were rated significantly higher in both GPT models compared to neurosurgeon experts suggests the importance of integrating domain-specific knowledge in the GPT models to improve response utility. Similarly, accuracy, relevance, and overall performance metrics were roughly equivalent between the GPT models and neurosurgeons. Interestingly, the GPT models were rated as having “minimal inaccuracies,” while the neurosurgeons were rated to have “some inaccuracies” to “minimal inaccuracies.”

Furthermore, the GPT models and neurosurgeons were rated as having “average relevance” to being “relatively relevant.” In the unblinded survey, there was a mix of responses across affective metrics, though the majority expressed positive opinions on GPT (Fig. 4). The relative superiority of GPT may relate to the impromptu nature of expert surgeon responses, i.e., as if they were speaking to a trainee and not at a board exam. Nonetheless, these results suggest that LLMs bear promise as a clinical aid.

### LLMs in the clinic

Clinical decision-making is a complex process that involves the application of practitioner knowledge and the integration of disease, patient, and system-level factors learned over time.^13,14^ Traditional methods of clinical decision-making can be hindered by limited access to relevant information, the need to integrate large amounts of complex information, and the possibility of human error, particularly in a fast-paced clinical environment.^13,14^ This may lead to variations in clinical practice amongst institutions, practitioners, sub-specialties (e.g., neurosurgery, otolaryngology, radiology, radiation oncology), and patient perceptions.^15–17^

The introduction of neuroGPT-X offers a unique tool for clinicians with rapid access to a wealth of information that includes patient-related findings and evidence-based guidelines, all integrated, summarized, and presented in a tailored and comprehensive manner.^18^ Such a tool can support decision-making by providing standardized processes and assisting practitioners with point-of-care informational aids.^16,19,20^ Interestingly and perhaps as expected, neuroGPT-X was rated as having more comprehensive responses than the neurosurgeons, whose answers reflected problems or nuances associated with the care of an individual patient rather than factual information that can be extracted from the literature. Nonetheless, clinicians retain and reference relevant textbook or literature information in their daily practice and, in particular, have knowledge reinforced by their clinical experiences.^21^ It is well known that human memory and recall are limited and can easily be overwhelmed while also being susceptible to biases and heuristics, which can influence information recall .^3,17, 22−24, 25^ Through rapid processing of current publications, neuroGPT-X provides clinicians with information that would be otherwise impossible to read or assimilate.

Furthermore, the neuroGPT-X framework is transferable between LLMs, allowing the potential for rapid advancements in the performance of AI-based medical decision-making systems. For example, integrating the framework with future iterations of BioGPT^26^ can benefit from improved clinical communications, as BioGPT was trained on millions of biomedical research articles to perform tasks such as answering questions, data extraction, and text generation. Indeed, as a framework with continual selective data input, neuroGPT-X enables the model and, by extension, its users, to adapt to incoming discoveries, guidelines, and evidence-based best practices.

### LLMs with a caution

Upon unblinding evaluation results and responses, the expert surgeons shared that the context-enriched GPT was “quite useful in clinical practice in the present age when physicians have to deal with huge amounts of data and knowledge” and that the answers “appear to be well backed up by evidence.” However, several salient concerns and questions were raised regarding the model’s limitations or interpretations (**Table 2**). The concerns, in part, may reflect a lack of level 1 evidence in managing vestibular schwannoma, with surgeons and sites opting for center-specific preferences, e.g., surgery versus radiosurgery versus observation. The GPT model relied primarily on available literature with possible biases. Indeed, if neuroGPT-X were to be advanced toward a reliable and clinically usable tool, relevant thematic breadth and depth of information would need to be incorporated. Moreover, the model must continually mature through an automated feed of filtered, selected, and complete (full paper) literature sources. Similarly, security and restrictions for online abuse, malware, or spam questioning to the model must be duly incorporated.

Although the ability of LLMs to exceed human performance in clinical acumen is promising, it is critical to ensure their responses are safe and reliable. LLMs are known to “hallucinate,” which refers to the generation of factually incorrect statements.^27,28^ More specifically, LLMs operate on the principle of “next best guess,” i.e., maximum likelihood estimation, based on probability distributions it learned via training. This raises questions about the nature of decision-making, both human and artificial. For example, human surgeons do not need a precise understanding of the trajectory of their joints when reaching out to grab a scalpel; they make a series of educated guesses based on their training and experience. Similarly, LLMs make estimates based on the probability distributions learned from the training data. However, LLMs cannot yet understand the implications of their decisions in the same way as humans. Therefore, it is essential to ensure that the responses provided by LLMs are accurate, safe, and reliable, a factor we attempted to address in neuroGPT-X through the inclusion of references and in-text citations.

### Human superiority over LLMs

Human memory is not solely storage and recall. It is also closely tied to experiences, emotions, intuition, and beliefs, all of which shape worldviews. This multifaceted nature of human memory may be challenging for LLMs to replicate. Moreover, human memory has advantages over LLMs in information processing and ideating. For example, humans routinely engage in creative thinking and problem-solving, enabling them to generate new ideas and innovative solutions. In contrast, LLMs are limited to answer generation based on their training dataset and the context supplied by their creator; their reliance on probability distributions also makes them more susceptible to inaccurate “hallucinations” or guessing.^27,28^ In this sense, LLMs fundamentally differ from human beings regarding their decision-making processes and abilities. These differences in memory, information processing, and recall necessitate the coupling of LLM technologies like neuroGPT-X with experienced practitioners to ensure safe and reliable practices in the clinical setting.

### Future directions

Many exciting possibilities exist for the ongoing improvement, evolution, and integration of LLMs in the clinical environment. A promising area is the development of multi-modal LLMs, which can interpret and integrate numerous inputs such as images, text, and other sensory data.^29^ Many medical specialties, including neurosurgery, rely heavily on multiple information modalities (e.g., imaging, patient demography, genetics, comorbidities, etc.) to arrive at well-reasoned decisions. Therefore, incorporating this information into decision-making can lead to a holistic care plan more attuned to a patient-specific clinical scenario. Continual growth and maturity of the model would also ensure up-to-date answers consistent with the latest standards of care. In particular, incorporating patient-related factors, such as preferences and values, into the model would further refine the reliability and inclusivity of the model.

With healthcare informatics, it is paramount to consider data security and confidentiality in applying LLMs to healthcare.^30^ The neuroGPT-X model hosted on a secure cloud platform already ensures security and compliance offered by Microsoft. From a systems-level perspective, a targeted effort to define the scope and policies surrounding LLMs in the healthcare setting must be considered as the platform becomes globally accessible.

### In conclusion

our study presents a responsive and clinically relevant AI tool, neuroGPT-X, utilizing context enrichment of the OpenAI GPT platform. The framework is readily transferable to other LLMs, opening possibilities for expanding its utility across neurosurgical diseases and other medical subspecialties. The study further highlights the importance of responsible AI development and the potential for machine intelligence to enhance clinical judgment while recognizing its limitations to ensure safe implementation into clinical practice.

## Data Availability

All data and code are available on Mendeley Data at this doi: 10.17632/b9mck42r35.1.

