## Supplementary Table 1 for "neuroGPT-X: Towards an Accountable Expert Opinion Tool for Vestibular Schwannoma"

**Supplementary Table 1.** Questions curated by a neurosurgeon with 30+ years of experience working with patients with vestibular schwannoma. Responses were given by ChatGPT, context-enriched GPT, neuroGPT-X, and the four expert neurosurgeons.

**Curated Questions**

1. What are the anatomical landmarks of the cerebellar-pontine angle?
2. What are the differential diagnoses of lesions within the cerebellar-pontine angle?
3. What is the age-specific incidence acoustic/vestibular schwannoma?
4. How do patients with acoustic/vestibular schwannoma present to their physicians?
5. How is acoustic/vestibular schwannoma diagnosed?
6. What are the imaging characteristics of vestibular schwannoma in computerized tomography (CT) and magnetic resonance (MR) imaging studies?
7. What are the treatment options? What are the indications for surgical treatment of acoustic/vestibular schwannoma?
8. What size of acoustic/vestibular schwannoma is amenable to radiosurgery?
9. What surgical approaches may be used in resecting acoustic/vestibular schwannoma, and why?
10. How does imaging characteristics and/or size influence choice of surgical approach in the excision of vestibular schwannoma?
11. What is the time course of radiation induced damage to the cochlear nerve following radiosurgery?
12. What are the risks associated with surgery versus radiosurgery for acoustic/vestibular schwannoma?
13. What are the treatment options for a patient with a 2cm right acoustic/vestibular schwannoma presenting with right sided tinnitus?
14. Is there a genetic pre-disposition for acoustic/vestibular schwannoma?
15. What is the management of a patient with bilateral acoustic/vestibular schwannoma, who presents with mild-moderate bilateral hearing loss?
