## Supplementary Table 2 for "neuroGPT-X: Towards an Accountable Expert Opinion Tool for Vestibular Schwannoma"

**Supplementary Table 2.** Evaluation criteria used by the expert neurosurgeons in the blinded and unblinded phases of evaluation.

| **Phase** | **Questions** | **Likert Scale Description** |
| --- | --- | --- |
| Evaluation (Blinded) | How accurate is this response? | 0 = completely inaccurate  1 = many inaccuracies  2 = some inaccuracies  3 = minimal inaccuracies  4 = completely accurate |
|  | How coherent is this response? | 0 = very incoherent and difficult to understand  1 = somewhat incoherent and not easily understood  2 = average coherence  3 = relatively coherent and understandable  4 = highly coherent and easy to understand |
|  | How relevant is this response? | 0 = not relevant at all  1 = somewhat relevant  2 = average relevance  3 = relatively relevant  4 = highly relevant |
|  | How thorough is this response? | 0 = not thorough at all  1 = somewhat thorough  2 = average thoroughness  3 = relatively thorough  4 = highly thorough |
|  | What is your overall rating of this response? | 0 = unusable in the clinic  1 = performs at the fellowship level  2 = performs at the attending level  3 = performs at the subspecialist level  4 = outperforms most subspecialists |
|  | Do you think this answer was written by a neurosurgeon or by GPT? | Neurosurgeon/GPT |
| Affective (Unblinded) | What is your overall satisfaction with this tool? | 0 = very dissatisfied  1 = dissatisfied  2 = neutral  3 = satisfied  4 = very satisfied |
|  | How likely would you use this tool in the clinic? | 0 = very unlikely  1 = unlikely  2 = neutral  3 = likely  4 = very likely |
|  | How much value do you perceive this tool to have? | 0 = no value  1 = low value, less than your current resources  2 = average value, on par with your current resources  3 = above average value, more than your current resources  4 = high value, significantly more than your current resources |
|  | How likely would you recommend this tool to your colleagues? | 0 = strongly unlikely  1 = unlikely  2 = neutral  3 = likely  4 = strongly likely |
| Long answer questions | What are your thoughts on this tool?  Do you have any other comments? | -- |
