## Supplementary Table 3 for "neuroGPT-X: Towards an Accountable Expert Opinion Tool for Vestibular Schwannoma"

**Supplementary Table 3.** Thematic analysis of each cluster from the vestibular schwannoma dataset using the InstructGPT Davinci model. Keyword summaries were made by instructing the InstructGPT Davinci model to summarize the thematic analysis results.

| **Cluster** | **Keywords** | **Thematic Analysis** |
| --- | --- | --- |
| 1 | Diseases, Diagnosis | All of the abstracts describe diseases and their diagnosis. |
| 2 | Vestibular Schwannoma, Treatments, Outcomes | The abstracts all discuss vestibular schwannoma and the various treatments and outcomes associated with it. |
| 3 | Vestibular Schwannoma, Neurofibromatosis, Pathophysiology, Treatment, Prognosis | The abstracts all discuss vestibular schwannoma, a type of tumor that can occur in patients with neurofibromatosis. They all discuss different aspects of the tumor, such as its pathophysiology, treatment, and prognosis. |
| 4 | Medical Care, Specialties, Hospitals, Degrees, Identifiers | The abstracts all discuss different aspects of medical care, including medical specialties, hospitals, degrees offered, and unique identifiers. |
| 5 | Vestibular Schwannoma, Diagnose, Treat, Quality of Life | The abstracts all discuss vestibular schwannoma, a type of tumor that affects the vestibular system. They also discuss various methods of diagnosing and treating the condition, as well as the impact it has on quality of life. |
| 6 | Vestibular Schwannoma, Tumor, Size, Growth Rate, Effects | The abstracts all discuss vestibular schwannoma, a type of tumor that affects the vestibular system. They all discuss different aspects of the tumor, such as its size, growth rate, and effects on the vestibular system. |
| 7 | Cancer, Study | The common theme among these abstracts is the study of cancer and its related topics. |
| 8 | Vestibular Schwannoma, Benign Tumor, Vestibulocochlear Nerve | All of the abstracts are related to vestibular schwannoma, a benign tumor of the vestibulocochlear nerve. |
| 9 | Nervous System, Functions | The abstracts all describe different parts of the nervous system and their functions. |
| 10 | Vestibular Schwannoma, Tumor, Facial Nerve | All of the abstracts are related to vestibular schwannoma, a type of tumor that affects the facial nerve. |
| 11 | Tumors, Treatments | The abstracts all discuss different types of tumors and treatments for them. |
