## Supplementary Table 4 for "neuroGPT-X: Towards an Accountable Expert Opinion Tool for Vestibular Schwannoma"

**Supplementary Table 4.** Questions curated by a neurosurgeon with 30+ years of experience working with patients with vestibular schwannoma. Responses given by ChatGPT, context-enriched GPT, neuroGPT-X, and the four expert neurosurgeons.

| Questions | 1. What are the anatomical landmarks of the cerebellar-pontine angle? 2. What are the differential diagnoses of lesions within the cerebellar-pontine angle? 3. What is the age-specific incidence acoustic/vestibular schwannoma? 4. How do patients with acoustic/vestibular schwannoma present to their physicians? 5. How is acoustic/vestibular schwannoma diagnosed? 6. What are the imaging characteristics of vestibular schwannoma in computerized tomography (CT) and magnetic resonance (MR) imaging studies? 7. What are the treatment options? What are the indications for surgical treatment of acoustic/vestibular schwannoma? 8. What size of acoustic/vestibular schwannoma is amenable to radiosurgery? 9. What surgical approaches may be used in resecting acoustic/vestibular schwannoma, and why? 10. How does imaging characteristics and/or size influence choice of surgical approach in the excision of vestibular schwannoma? 11. What is the time course of radiation induced damage to the chochlear nerve following radiosurgery? 12. What are the risks associated with surgery versus radiosurgery for acoustic/vestibular schwannoma? 13. What are the treatment options for a patient with a 2cm right acoustic/vestibular schwannoma presenting with right sided tinnitus? 14. Is there a genetic pre-disposition for acoustic/vestibular schwannoma? 15. What is the management of a patient with bilateral acoustic/vestibular schwannoma, who presents with mild-moderate bilateral hearing loss? |
| --- | --- |
| ChatGPT | 1. The cerebellopontine angle (CPA) is a triangular region located between the cerebellum and the pons, and contains several key anatomical landmarks. These landmarks include:  1. The internal auditory meatus (IAM): a bony canal in the temporal bone that houses the vestibulocochlear nerve (CN VIII), responsible for hearing and balance. 2. The facial nerve (CN VII): responsible for motor control of the face and associated muscles. 3. The vestibular nerve: responsible for transmitting information about balance and head position to the brain. 4. The glossopharyngeal nerve (CN IX): responsible for sensory and motor control of the tongue, pharynx, and larynx. 5. The vagus nerve (CN X): responsible for sensory and motor control of the viscera, including the heart, lungs, and digestive system. 6. The anterior inferior cerebellar artery (AICA): one of the main blood supplies to the brainstem and inner ear.  The precise definition of the CPA and its landmarks can vary between sources, but these structures are commonly considered key landmarks in this region. An understanding of the anatomy of the CPA is important for various medical procedures, including the removal of tumors and the placement of electrodes for functional neuromodulation. 2. Lesions within the cerebellopontine angle (CPA) can present a range of clinical signs and symptoms, and a wide variety of differential diagnoses should be considered in the evaluation of such lesions. Some of the differential diagnoses include:  1. Vestibular schwannoma (acoustic neuroma): a slow-growing, benign tumor of the vestibular nerve, often presenting with hearing loss and tinnitus. 2. Meningioma: a slow-growing, benign tumor of the meninges, often presenting with cranial nerve symptoms and hearing loss. 3. Epidermoid cyst: a benign cyst derived from ectodermal tissue, often presenting with hearing loss and tinnitus. 4. Arachnoid cyst: a benign cyst in the subarachnoid space, often presenting with cranial nerve symptoms and hydrocephalus. 5. Trigeminal schwannoma (trigeminal neuralgia): a slow-growing, benign tumor of the trigeminal nerve, often presenting with facial pain and numbness. 6. Metastatic disease: malignant tumors that have spread to the CPA from elsewhere in the body, often presenting with cranial nerve symptoms and rapidly progressive neurologic deficit. 7. Multiple sclerosis: an autoimmune disease affecting the central nervous system, often presenting with cranial nerve symptoms and hearing loss. 8. The specific differential diagnoses will depend on the patient's symptoms and the findings on imaging studies, and the final diagnosis may require biopsy or further evaluation. An accurate diagnosis is crucial for appropriate treatment planning and prognostication. 3. Vestibular schwannomas, also known as acoustic neuromas, are tumors that develop from the cells that surround the vestibular nerve. The age-specific incidence of vestibular schwannomas varies, with the majority of cases occurring in people over the age of 50. The age-specific incidence of vestibular schwannomas is as follows:  1. 50-60 years: Low incidence rate, with a gradual increase over time. 2. 60-70 years: The incidence rate begins to increase significantly. 3. 70-80 years: The incidence rate reaches its peak, with the majority of cases diagnosed in this age range. 4. Over 80 years: The incidence rate begins to decrease again, but there is still a significant number of cases in this age range.  It is important to note that vestibular schwannomas can occur in people of any age, but they are more common in people over the age of 50. In addition, certain genetic conditions, such as neurofibromatosis type 2, can increase the risk of developing a vestibular schwannoma at a younger age. 4. Patients with vestibular schwannoma (acoustic neuroma) may present to their physician with a variety of symptoms, depending on the size and location of the tumor. Common presenting symptoms include:  1. Hearing loss: The most common symptom, usually starting in one ear and affecting both high and low frequencies. 2. Tinnitus: Ringing or buzzing in the affected ear. 3. Dizziness or unsteadiness: Due to involvement of the vestibular nerve. 4. Facial weakness or numbness: Due to compression of the facial nerve. 5. Headache: Rare but can occur if the tumor is large and compresses surrounding structures. 6. Balance problems: Due to involvement of the vestibular nerve. 7. Cognitive or memory changes: Rare but can occur if the tumor is large and compresses surrounding structures.  The specific symptoms will depend on the size and location of the tumor, as well as the patient's overall health. Some patients may have few or no symptoms, while others may experience a wide range of symptoms that affect their daily life. An accurate diagnosis is crucial for appropriate treatment planning and prognostication, and may require imaging studies such as magnetic resonance imaging (MRI) or computed tomography (CT) scans. 5. Diagnosis of vestibular schwannoma (acoustic neuroma) typically begins with a thorough history and physical examination, including a review of the patient's symptoms and any relevant medical history. Other diagnostic tools and tests that may be used to diagnose vestibular schwannoma include:  1. Audiological evaluation: An evaluation of the patient's hearing, including pure-tone audiometry and speech audiometry, to assess for hearing loss in specific frequencies. 2. Imaging studies: Magnetic resonance imaging (MRI) is the preferred imaging modality for vestibular schwannomas, as it provides clear images of the tumor and surrounding structures. Computed tomography (CT) scans may also be used to help diagnose vestibular schwannomas. 3. Vestibular function tests: Tests to assess the function of the vestibular nerve, including caloric testing, rotational testing, and vestibular evoked myogenic potential (VEMP) testing. 4. Biopsy: In rare cases, a biopsy of the tumor may be necessary to confirm the diagnosis and exclude other types of tumors.  The combination of the patient's symptoms, medical history, and results from the above tests will help the physician determine the diagnosis of vestibular schwannoma and plan appropriate treatment. It is important to have a thorough evaluation and an accurate diagnosis to ensure the best possible outcome. 6. Vestibular schwannomas, also known as acoustic neuromas, can be visualized on both computerized tomography (CT) and magnetic resonance (MR) imaging studies. The imaging characteristics of vestibular schwannomas on these studies include:  CT:  1. Isodense or hypodense on non-contrast CT, compared to brain parenchyma. 2. Well-defined borders, with homogenous density within the tumor. 3. Typically not enhancing after contrast administration, although peripheral enhancement may occur. 4. May demonstrate erosion of adjacent bony structures such as the internal auditory canal.  MR:  1. Hypointense on T1-weighted images, and hyperintense on T2-weighted images. 2. Typically well-defined margins, with homogenous signal intensity within the tumor. 3. May enhance after contrast administration, with peripheral enhancement more common than central enhancement. 4. May demonstrate invasion of adjacent brainstem structures.  It is important to note that the imaging characteristics of vestibular schwannomas may vary depending on the size, location, and stage of the tumor. In addition, other types of tumors can present with similar imaging features, so a thorough evaluation of the patient's symptoms and medical history is also important for accurate diagnosis. 7. The treatment options for vestibular schwannoma (acoustic neuroma) include:  1. Observation: For small tumors or tumors that are not causing significant symptoms, observation may be a suitable option. The patient will be monitored with regular imaging studies to assess for tumor growth. 2. Radiation therapy: This may include stereotactic radiosurgery (SRS) or fractionated radiotherapy. Radiation therapy can shrink the tumor and help control symptoms, but is typically not curative. 3. Surgical resection: This involves removing the tumor through an incision in the skull. The surgical approach will depend on the size and location of the tumor, as well as the patient's overall health and symptoms. The most common surgical approach for vestibular schwannomas is the retrosigmoid approach.  The indications for surgical treatment of vestibular schwannoma include:  1. Tumor growth: If the tumor is growing and causing significant symptoms, surgery may be indicated. 2. Progressive hearing loss: If the tumor is causing progressive hearing loss, surgery may be indicated to preserve the remaining hearing. 3. Involvement of adjacent cranial nerves: If the tumor is causing weakness or numbness in the face, or other symptoms related to involvement of adjacent cranial nerves, surgery may be indicated. 4. Large tumors: For large tumors, surgery may be the only option to control symptoms and prevent further damage to adjacent structures.  The treatment approach for vestibular schwannoma will depend on several factors, including the size and location of the tumor, the patient's overall health, and the patient's symptoms. A team of specialists, including a neurosurgeon and a neuro-otologist, will work together to determine the best course of treatment for each individual patient. 8. The size of a vestibular schwannoma that is amenable to radiosurgery can vary depending on several factors, including the patient's overall health and the location of the tumor. In general, radiosurgery is typically considered a viable treatment option for tumors that are less than 4 cm in diameter.  It is important to note that not all patients with vestibular schwannomas are suitable candidates for radiosurgery, and that the optimal treatment approach will depend on several factors, including the size and location of the tumor, the patient's overall health, and the patient's symptoms. A team of specialists, including a neurosurgeon and a neuro-otologist, will work together to determine the best course of treatment for each individual patient.  It is also important to note that the long-term effectiveness and safety of radiosurgery for vestibular schwannomas is still being studied, and that patients should be carefully monitored after undergoing radiosurgery to assess for any potential adverse effects. 9. There are several surgical approaches that may be used in resecting vestibular schwannomas (acoustic neuromas), including:  1. Translabyrinthine approach: This approach involves accessing the tumor through an incision behind the ear. This approach is typically used for larger tumors or tumors that are located more deeply within the skull. 2. Retrosigmoid approach: This approach involves accessing the tumor through an incision at the base of the skull, in the back of the head. This approach is less invasive than the translabyrinthine approach and is typically used for smaller tumors or tumors that are located more superficially. 3. Middle cranial fossa approach: This approach involves accessing the tumor through an incision in the skull, above the ear. This approach is typically used for tumors that are located in the middle cranial fossa, and is less commonly used than the other two approaches.  The choice of surgical approach will depend on several factors, including the size and location of the tumor, the patient's overall health, and the patient's symptoms. A team of specialists, including a neurosurgeon and a neuro-otologist, will work together to determine the best surgical approach for each individual patient.  In general, the goal of surgical treatment of vestibular schwannomas is to remove as much of the tumor as possible while preserving the patient's hearing and neurological function. The specific surgical approach used will depend on the location and size of the tumor, as well as the patient's overall health and symptoms. 10. The imaging characteristics and size of a vestibular schwannoma (acoustic neuroma) can influence the choice of surgical approach in its excision. The following factors are usually considered when choosing a surgical approach:  1. Tumor size: Larger tumors may require a more extensive surgical approach, such as the translabyrinthine approach, to ensure complete removal. Smaller tumors may be amenable to a less invasive surgical approach, such as the retrosigmoid approach. 2. Location of the tumor: The location of the tumor within the skull can impact the choice of surgical approach. Tumors located in the middle cranial fossa may require a different surgical approach compared to those located more superficially or deeper within the skull. 3. Imaging characteristics: The imaging characteristics of the tumor, such as its shape, consistency, and relationship to surrounding structures, can also influence the choice of surgical approach. For example, tumors that are closely associated with critical structures, such as the facial nerve or the brainstem, may require a more specialized surgical approach to minimize the risk of injury to these structures. 4. Patient's hearing status: The patient's hearing status can also impact the choice of surgical approach. Patients with residual hearing may be more likely to benefit from a less invasive surgical approach, such as the retrosigmoid approach, as this approach may preserve their hearing to a greater extent.  Ultimately, the choice of surgical approach will depend on the individual patient and the specific characteristics of their vestibular schwannoma. A team of specialists, including a neurosurgeon and a neuro-otologist, will work together to determine the best surgical approach for each individual patient. 11. The time course of radiation-induced damage to the cochlear nerve following radiosurgery can vary, but it typically occurs gradually over a period of months to years. The exact timing and severity of the damage will depend on several factors, including the dose of radiation delivered to the nerve, the patient's age, and their overall health.  In some cases, patients may experience a decline in their hearing soon after radiosurgery. However, for many patients, the effects of radiation on the cochlear nerve may not become apparent until several months or years later. In some cases, the damage may continue to progress over time, leading to a gradual decline in hearing.  It is important to note that not all patients who undergo radiosurgery for a vestibular schwannoma (acoustic neuroma) will experience radiation-induced damage to the cochlear nerve. The risk of damage will depend on several factors, including the dose and location of the radiation, as well as the patient's overall health and age. A team of specialists, including a neurosurgeon and a neuro-otologist, will work together to determine the best treatment plan for each individual patient. 12. Both surgery and radiosurgery for vestibular schwannoma (acoustic neuroma) are associated with risks and benefits, and the choice of treatment will depend on several factors, including the size of the tumor, the patient's age, and overall health, and the patient's hearing status.  Risks associated with surgical excision of vestibular schwannoma include:  1. Hearing loss: Surgical removal of a vestibular schwannoma can result in permanent hearing loss in the affected ear. 2. Facial nerve paralysis: The facial nerve runs close to the vestibular schwannoma, and injury to this nerve can occur during surgery, resulting in temporary or permanent facial paralysis. 3. CSF (cerebrospinal fluid) leak: Surgery in the area of the vestibular schwannoma can result in a leak of cerebrospinal fluid (CSF), which can cause headaches, meningitis, or other complications. 4. Brain injury: There is a risk of injury to the brain or other critical structures during surgery.  Risks associated with radiosurgery for vestibular schwannoma include:  1. Radiation injury to the cochlear nerve: Radiosurgery for vestibular schwannoma can result in injury to the cochlear nerve, leading to progressive hearing loss over time. 2. Tumor regrowth: In some cases, the vestibular schwannoma may regrow after radiosurgery, requiring additional treatment. 3. Cognitive decline: There is a theoretical risk of cognitive decline after radiosurgery, although the evidence for this is limited and the risk is believed to be low.  It is important to discuss the risks and benefits of both surgical excision and radiosurgery with a neurosurgeon and a neuro-otologist to determine the best treatment plan for each individual patient. 13. The treatment options for a patient with a 2 cm right-sided vestibular schwannoma (acoustic neuroma) presenting with right-sided tinnitus will depend on several factors, including the patient's age, overall health, and hearing status. In general, the options include:  1. Observation: For small tumors that are not causing symptoms, it may be appropriate to simply observe the tumor and monitor its growth over time. 2. Radiosurgery: For patients who are not good candidates for surgery or who prefer a minimally invasive option, radiosurgery may be an option. This treatment uses high-dose radiation to shrink the tumor and is typically performed as an outpatient procedure. 3. Microsurgical excision: Microsurgical excision is the most common treatment for vestibular schwannoma and involves removing the tumor through a small incision in the skull. This approach provides the best chance of preserving hearing and other cranial nerve function.  In the case of a 2 cm vestibular schwannoma, both observation and radiosurgery may be reasonable options for some patients, while microsurgical excision may be the preferred choice for others. The patient's physician, in collaboration with a neurosurgeon and a neuro-otologist, will determine the best treatment plan for the individual patient based on their specific needs and circumstances. 14. Yes, there is a genetic predisposition for acoustic/vestibular schwannoma, also known as an acoustic neuroma. About 10-15% of all cases of acoustic neuroma are associated with genetic conditions such as neurofibromatosis type 2 (NF2) and hereditary acoustic neuroma.  NF2 is an autosomal dominant disorder that is associated with the development of multiple tumors of the nervous system, including bilateral acoustic neuromas in over 50% of cases. In hereditary acoustic neuroma, the tumor occurs in families with a history of the condition and is caused by a mutation in the NF2 gene.  Having a family history of acoustic neuroma or a known genetic condition such as NF2 may increase an individual's risk for developing the condition. However, most cases of acoustic neuroma are not associated with a known genetic predisposition and occur sporadically. 15. The management of a patient with bilateral acoustic/vestibular schwannoma (BNAs) and mild-moderate bilateral hearing loss will depend on several factors, including the size and growth rate of the tumors, the patient's overall health, and their individual preferences and goals.  In general, the following options may be considered:  1. Observation: For small tumors that are not causing significant symptoms, observation with regular imaging may be appropriate. 2. Radiosurgery: Radiosurgery is a minimally invasive option that can be used to shrink tumors and slow or stop their growth. This option is typically reserved for smaller tumors or for patients who are not good candidates for surgery. 3. Microsurgical excision: Microsurgical excision involves removing the tumors through small incisions in the skull. This approach can provide the best chance of preserving hearing and other cranial nerve function, but also carries a risk of complications, including facial weakness and hearing loss. 4. Hearing aids or cochlear implants: In some cases, hearing aids or cochlear implants may be recommended to help the patient manage their hearing loss, regardless of the treatment choice for their tumors.  The patient's physician, in collaboration with a neurosurgeon and a neuro-otologist, will determine the best treatment plan for the individual patient based on their specific needs and circumstances. In some cases, a multidisciplinary team approach may be necessary to manage the complex needs of patients with BNAs. |
| Context-Enriched GPT | 1. The cerebellopontine angle (CPA) is located between the cerebellum and the pons. It is bound anterolaterally by the posterior aspect of the petrous temporal bone and posteromedially by the cerebellum and pons. It contains important vascular structures and cranial nerves and is subject to a certain gamut of lesions, notably tumors with interesting radiological manifestations. The cerebellopontine angle is formed by the cerebellopontine fissure, which is made when the cerebellum folds over to the pons, creating a sharply defined angle between them. The angle formed in turn creates a subarachnoid cistern, the cerebellopontine angle cistern. The pia mater follows the outline of the fissure and the arachnoid mater continues across the divide so that the subarachnoid space is dilated at this area, forming the cerebellopontine angle cistern. The anterior inferior cerebellar artery (AICA) is the principal vessel of the cerebellopontine angle. It also contains two cranial nerves – the vestibulocochlear nerve and the facial nerve; the cerebellar flocculus and the lateral recess of the fourth ventricle. 2. The differential diagnoses of lesions within the cerebellopontine angle include vestibular schwannoma, meningioma, epidermoid cyst, lipoma, metastases, glioma, ependymoma, choroid plexus papilloma, and vascular malformations. Other rarer lesions include arachnoid cyst, facial nerve tumour, schwannoma of other cranial nerves, intracranial epidermoid cyst, and cerebellar astrocytoma. 3. The age-specific incidence of acoustic/vestibular schwannoma varies depending on the population studied. According to a study of the incidence of vestibular schwannoma in Taiwan, the highest incidence rate of 4.86 per 100,000 persons was observed in the age group of 60 to 69 years. In a study of the incidence of vestibular schwannoma in Denmark, the age-specific incidence rates show that VS is rarely diagnosed in persons under 30, but the incidence rate appears to increase among women after menopause. In a study of the incidence of vestibular schwannoma in the United States, incidence increased with age to a peak of 2.93 per 100,000 in the 65-74 year old age group. 4. Patients with acoustic/vestibular schwannoma typically present to their physicians with unilateral or asymmetric hearing loss, which is the most common initial symptom. Other symptoms may include tinnitus, feeling of insecurity or vertigo, cranial nerve palsies, and headache. In patients with preexisting cognitive impairment, the most common reasons for seeking medical care include worsening ataxia and headaches, despite the fact that all patients had profound ipsilateral hearing loss. Additionally, some patients may endorse facial numbness, and partial facial paresis. 5. Acoustic/vestibular schwannoma is typically diagnosed using magnetic resonance imaging (MRI). MRI is the gold standard for diagnosing this type of tumor due to its high sensitivity and specificity. Other diagnostic tests that may be used to diagnose acoustic/vestibular schwannoma include audiologic diagnostics such as supraliminary audiometry, tympanometry, stapedius reflex and otoacoustic emissions, as well as vestibular response to caloric testing. Brainstem Auditory Evoked Potentials (BAEP) is the golden standard for audiologic diagnostics of vestibular schwannoma. If pathological findings of BAEP are present, then an MRI of the posterior fossa should be performed. Gadolinium-enhanced MRI is the best and final tool for making a diagnosis of vestibular schwannoma. 6. Vestibular schwannomas are typically seen as a well-defined, homogeneous, and hyperintense mass on T1-weighted MR images. On T2-weighted images, the mass is hypointense. On contrast-enhanced T1-weighted images, the mass is typically hyperintense and shows homogeneous enhancement. On CT scans, the mass is typically seen as a hypodense mass with a thin rim of enhancement. The mass may also show calcification or cystic changes. 7. The treatment options for acoustic/vestibular schwannoma include observation, open surgical resection, stereotactic radiosurgery, fractionated radiotherapy, and medical therapy. The indications for surgical treatment of acoustic/vestibular schwannoma include large tumors causing mass effect and obstructive hydrocephalus, tumors that are growing rapidly, and tumors that are causing significant symptoms. Surgery is also indicated for tumors that are not amenable to radiosurgery or radiotherapy, or for tumors that are located in areas that are difficult to access with radiosurgery or radiotherapy. 8. Radiosurgery is an effective treatment option for vestibular schwannomas of appropriate size, typically smaller than 4 cm3. Studies have shown that radiosurgery can arrest tumor growth in up to 98% of patients when studied at intervals of 10-15 years. In addition, radiosurgery has a high safety level, using low radiation doses, low rate of complications, and good tumor control rate in the medium term follow-up. 9. The three main surgical approaches used in resecting acoustic/vestibular schwannoma are the translabyrinthine, retrosigmoid, and subtemporal microsurgical approaches. The translabyrinthine approach is used when the tumor is large and/or when the patient has pre-existing neurologic deficits. This approach involves removing the entire inner ear and facial nerve, and is the most invasive of the three approaches. The retrosigmoid approach is used when the tumor is smaller and/or when the patient has pre-existing hearing. This approach involves removing the tumor through the posterior fossa and preserving the facial nerve. The subtemporal approach is used when the tumor is small and/or when the patient has pre-existing hearing. This approach involves removing the tumor through the temporal bone and preserving the facial nerve. The choice of approach depends on the size of the tumor, pre-existing neurologic deficits, and the patient's hearing status. 10. Imaging characteristics and size are important factors in determining the optimal surgical approach for the excision of vestibular schwannoma. Magnetic resonance imaging (MRI) is the imaging method of choice for patients with vestibular schwannoma, with almost 100% sensitivity and specificity. High-resolution sequences and the application of gadolinium-based i.v. contrast medium are crucial for MRI-based diagnosis. Description of the size and location of the tumor, as well as its relationship to adjacent structures and anatomic features, is essential, since these factors, in addition to clinical findings, influence the type of treatment, surgical options, and associated risks.  Smaller tumors (<2 cm) can be managed with a minimally invasive endoscopic approach, while larger tumors (>2 cm) may require a more extensive open approach. The size of the tumor is also important in determining the extent of the resection. For tumors larger than 2 cm, a gross total resection (GTR) is recommended, while for smaller tumors, a subtotal resection may be sufficient. The presence of tumor extension into the internal auditory canal or cerebellopontine angle can also influence the choice of surgical approach. In these cases, a more extensive open approach may be necessary to ensure complete tumor removal. 11. The time course of radiation induced damage to the cochlear nerve following radiosurgery is variable and depends on the radiation dose received by the cochlea. Generally, the higher the radiation dose, the more rapid the onset of hearing loss. In a study of 33 patients with vestibular schwannoma treated with gamma knife radiosurgery, the mean pretreatment pure-tone average (PTA) and speech discrimination score (SDS) were 55.86 dB and 45.70%, respectively. The mean PTA and SDS at last follow-up were 66.55 dB and 39.15%, respectively. The PTA at 6 months (p = 0.003), 12 months (p = 0.004), and last follow-up (p = 0.001) was significantly poorer than the pretreatment PTA. There was no significant difference between pretreatment and follow-up SDS at any time interval. The mean cochlear radiation dose was 5.2 Gy (range, 2.6-8.5 Gy). The median cochlear dose was 4.75 Gy. Fifteen patients received less than the median cochlear dose, and 18 received greater than or equal to the median cochlear dose. The change in PTA from baseline was significantly poorer at 12 months for those patients whose cochlea received 4.75 Gy (p = 0.02) or greater. This suggests that radiation induced damage to the cochlear nerve can occur as early as 6 months after radiosurgery, and that the risk of hearing loss increases with higher radiation doses. 12. Surgery for acoustic/vestibular schwannoma carries a risk of complications that are inherent to general anesthesia and surgery of any type, including myocardial infarction, pneumonia, pulmonary embolism, and infection. Additionally, there is a risk of specific complications related to neurosurgery in this area of the brain, such as hydrocephalus, cerebrospinal fluid leak, facial nerve paralysis, facial numbness, hearing loss, ataxia, dysphagia, and major stroke. Radiosurgery provides an outpatient, noninvasive alternative for the treatment of small acoustic schwannomas, but carries its own risks. A systematic review and meta-analysis comparing the outcomes of surgery versus stereotactic radiosurgery (SRS) found that both surgery and SRS resulted in similar odds of hearing loss and similar improvements in dizziness and tinnitus among patients with vestibular schwannoma; however, facial symmetry loss appeared higher post-surgery. Additionally, tumor control was larger in the surgery (94%) than the SRS group (80%) for small-to-medium size tumors. With fractionated radiotherapy, 3-7% will require additional treatment and hearing preservation is reported at 59-94% of patients, although long-term outcomes are not known. Microsurgery is an alternative for eligible patients, with fewer than 2% requiring additional treatment; however, the risk of hearing loss, facial neuropathy, and other morbidities is relatively high. 13. The treatment options for a patient with a 2cm right acoustic/vestibular schwannoma presenting with right sided tinnitus include observation, open surgical resection, stereotactic radiosurgery, and fractionated radiotherapy. Observation is a reasonable management option for small tumors that are not growing. Open surgical resection is an option for tumors that are larger than 2 cm and can be performed with the goal of preserving hearing. Stereotactic radiosurgery is an option for tumors that are smaller than 2.5 cm and have documented growth. Fractionated radiotherapy is an option for tumors that are larger than 2 cm and can be used to preserve hearing. The decision of which treatment option to pursue should be made on a case-by-case basis in consultation with a multidisciplinary team. 14. Yes, there is a genetic pre-disposition for acoustic/vestibular schwannoma. Studies have shown that mutations in the NF2 gene, which is responsible for the tumor suppressor protein merlin, are associated with an increased risk of developing vestibular schwannoma. Additionally, mutations in the LZTR1 gene have been linked to an increased risk of developing schwannomatosis, which is a condition characterized by multiple schwannomas. Furthermore, a genome-wide association study has identified a risk locus on chromosome 9p21.3, which is associated with an increased risk of developing vestibular schwannoma. This locus contains the CDKN2B-AS1 and CDKN2A/B genes, which are known to be involved in tumorigenesis. Finally, epigenetic events such as DNA methylation and histone modifications have been linked to the development and progression of hearing loss in NF2 patients. 15. The management of a patient with bilateral acoustic/vestibular schwannoma and mild-moderate bilateral hearing loss depends on the size of the tumors and the patient's age and overall health. If the tumors are small and the patient is young and healthy, then surgery may be recommended to remove the tumors and preserve hearing. Surgery may involve a translabyrinthine, retrosigmoid, or middle cranial fossa approach, depending on the size and location of the tumors. If the tumors are large or the patient is elderly or has other medical conditions, then a more conservative approach may be recommended, such as observation with regular MRI scans and audiometry. In this case, hearing aids may be recommended to help improve hearing. If the tumors grow or hearing deteriorates, then surgery may be recommended. |
| neuroGPT-X | 1. The anatomical landmarks of the cerebellar-pontine angle include the posterior aspect of the petrous temporal bone anterolaterally, the cerebellum and pons posteromedially, the cerebellopontine fissure, the cerebellopontine angle cistern, the anterior inferior cerebellar artery (AICA), the vestibulocochlear nerve, the facial nerve, the cerebellar flocculus, the lateral recess of the fourth ventricle, and the subarachnoid space [1, 3]. Additionally, the cerebellopontine angle is subject to a certain gamut of lesions, notably tumors with interesting radiological manifestations [2]. The most frequent lesion identified at the cerebellopontine angle cistern and internal auditory meatus (IAM) is the vestibular schwannoma [6], followed by meningioma and epidermoid cyst [4, 5]. Other lesions such as lipoma, papilloma, glioma or metastasis account for less than 1% of the cerebellopontine angle cistern tumors in adults [8]. The retrosigmoid craniotomy is the primary means by which to gain surgical access to this anatomically complex region [7].  Sources:  [1] Cerebellopontine angle, Summary, Wikipedia. [2] Vestibular schwannoma. Cerebellopontine angle tumors: role of magnetic resonance imaging., A A Zamani. [3] Cerebellopontine angle, Structure, Wikipedia. [4] Cerebellopontine angle syndrome, Summary, Wikipedia. [5] Vestibular schwannoma. Pathology of the cerebellopontine angle.J-L Sarrazin, K Marsot-Dupuch, A Chaÿas. [6] Vestibular schwannoma. The role of radiology in the diagnosis and management of vestibular schwannoma., N Sriskandan, S E J Connor. [7] Vestibular schwannoma. Left-sided retrosigmoid craniotomy for the resection of a vestibular schwannoma., Peter S Amenta, Jacques J Morcos. [8] Vestibular schwannoma. Cerebellopontine angle tumors in adults.J Sarrazin, O Hélie, Y Cordoliani. 2. The differential diagnoses of lesions within the cerebellar-pontine angle include vestibular schwannoma [1], meningioma [2], epidermoid cyst [4], lipoma [4], metastases [4], gliomas [2], ependymomas [2], choroid plexus papillomas [2], and vascular malformations [2]. Additionally, the presence of any suspicious clinical and radiographic findings uncharacteristic of vestibular schwannoma [6] may indicate the presence of other lesions such as arachnoid cyst [7], facial nerve tumour [7], schwannoma of other cranial nerves [7], or intracranial epidermoid cyst [7].  Sources:  [1] Vestibular schwannoma. The role of radiology in the diagnosis and management of vestibular schwannoma., N Sriskandan, S E J Connor. [2] Vestibular schwannoma. Cerebellopontine angle tumors: role of magnetic resonance imaging., A A Zamani. [3] Cerebellopontine angle syndrome, Summary, Wikipedia. [4] Vestibular schwannoma. Cerebellopontine angle tumors in adults.J Sarrazin, O Hélie, Y Cordoliani. [5] Vestibular schwannoma. Pathology of the cerebellopontine angle.J-L Sarrazin, K Marsot-Dupuch, A Chaÿas. [6] Vestibular schwannoma. Diagnostic and surgical challenges in resection of cerebellar angle tumors and acoustic neuromas., Neal Patel, Jared Wilkinson, Nicholas Gianaris, Aaron A Cohen-Gadol. [7] Cerebellopontine angle, Clinical significance, Wikipedia. 3. The age-specific incidence of acoustic/vestibular schwannoma varies depending on the population studied. According to a study of the Taiwanese population [1], the highest incidence rate of 4.86 per 100 000 persons was observed in the age group of 60 to 69 years. In a study of the Danish population [2], the overall incidence rate of vestibular schwannoma was found to be 5 cases per million population per year in 1977-1981, increasing to 10 cases in 1992-1995. In a study of the unselected population [3], the incidence of diagnosed sporadic unilateral vestibular schwannoma increased from 3.1 diagnosed VS per million per year in 1976 to a peak of 22.8 VS per million per year in 2004, which was followed by a decrease to 19.4 VS per million per year in 2008. Finally, a study of pediatric cases [4] found that the mean tumor size at diagnosis decreased from 30 mm in 1979 to 10 mm in 2008, whereas hearing acuity at diagnosis has improved over the years.  Sources:  [1] Vestibular schwannoma. Incidence of Vestibular Schwannoma in Taiwan from 2001 to 2012: A Population-Based National Health Insurance Study., Malcolm Koo, Jen-Tsung Lai, Edward Yih-Liang Yang, Tien-Chen Liu, Juen-Haur Hwang. [2] Vestibular schwannoma. Incidence of vestibular schwannoma in Denmark, 1977-1995., M F Howitz, C Johansen, M Tos, S Charabi, J H Olsen. [3] Vestibular schwannoma. True incidence of vestibular schwannoma?, Sven-Eric Stangerup, Mirko Tos, Jens Thomsen, Per Caye-Thomasen. [4] Vestibular schwannoma. Management of Sporadic Vestibular Schwannomas in Children-Volumetric Analysis and Clinical Outcome Assessment., Julian Zipfel, Mykola Gorbachuk, Isabel Gugel, Marcos Tatagiba, Martin U Schuhmann. 4. Patients with acoustic/vestibular schwannoma typically present to their physicians with unilateral hearing loss and tinnitus, as these are the most common symptoms associated with the condition [1]. Other symptoms that may be present include dizziness, facial paresthesias, headaches, vision changes, and ataxia [2, 4]. Patients with preexisting cognitive impairment may be unable to acknowledge these symptoms or communicate their presence, leading to a delay in diagnosis and treatment [4].  Sources:  [1] Vestibular schwannoma. Unusual clinical presentations of vestibular schwannomas.Andrés Coca Pelaz, Juan P Rodrigo, José L Llorente, Justo R Gómez, Carlos Suárez. [2] Vestibular schwannoma. Case series of vestibular schwannoma patients with no asymmetry in hearing., Brian Kellermeyer, Erica Haught, Tyler Harper, Stephen Wetmore. [3] Vestibular schwannoma. Main Symptom that Led to Medical Evaluation and Diagnosis of Vestibular Schwannoma and Patient-Reported Tumor Size: Cross-sectional Study in 1,304 Patients., Maria Peris-Celda, Christopher S Graffeo, Avital Perry, Panagiotis Kerezoudis, Nicole M Tombers, Matthew L Carlson, Michael J Link. [4] Vestibular schwannoma. Cognitive delay as a risk factor for late presentation of vestibular schwannoma., Matthew L Carlson, George B Wanna, Colin L Driscoll, Kyle D Weaver, Michael J Link. 5. Vestibular schwannoma is typically diagnosed using magnetic resonance imaging (MRI) [1]. MRI is the gold standard for diagnosing this condition, as it has a sensitivity and specificity of up to 100% [3]. Audiologic diagnostics, such as supraliminary audiometry, tympanometry, stapedius reflex, otoacoustic emissions, and vestibular response to caloric testing, are also used to select patients with suspicion of this tumor [4]. Brainstem Auditory Evoked Potentials (BAEP) is the golden standard for audiologic diagnostics of vestibular schwannoma [4]. If pathological findings of BAEP are present, then an MRI of the posterior fossa should be performed [4]. Additionally, biomarkers such as proteins that regulate merlin conformation changes, inflammatory cytokines, miRNAs, tissue proteins, and cerebrospinal fluid (CSF) components have been proposed to be closely related to tumor volume increase [5]. These biomarkers may be used to determine the optimal treatment strategy for each patient [5].  Sources:  [1] Vestibular schwannoma. Update on diagnostics and microsurgical treatment of vestibular schwannoma.F H Ebner, M Tatagiba. [2] Vestibular schwannoma. Quality of life in patients with vestibular schwannomas managed conservatively., C MacAndie, J A Crowther. [3] Vestibular schwannoma. Vestibular schwannoma: Part I: epidemiology and diagnostics.F Hassepass, S B Bulla, A Aschendorff, W Maier. [4] Vestibular schwannoma. Audiologic diagnostics of vestibular schwannoma.Zoran Komazec, Slobodanka Lemajić, Ljilana Vlaski. [5] Vestibular schwannoma. Potential Molecular Biomarkers of Vestibular Schwannoma Growth: Progress and Prospects., Yu Zhang, Jianfei Long, Junwei Ren, Xiang Huang, Ping Zhong, Bin Wang. [6] Vestibular schwannoma. Perioperative Care of the Patient With Acoustic Neuroma., Kristen A Oster. [7] Vestibular schwannoma. Facial nerve schwannoma: A case report and review of the literature., Mei-Chien Chen, Te-Ming Tseng, Shih-Han Hung, Po-Yueh Chen. [8] Vestibular schwannoma. EAONO Position Statement on Vestibular Schwannoma: Imaging Assessment. What are the Indications for Performing a Screening MRI Scan for a Potential Vestibular Schwannoma?, Jérôme Waterval, Romain Kania, Thomas Somers. 6. The imaging characteristics of vestibular schwannoma in computerized tomography (CT) and magnetic resonance (MR) imaging studies are typically seen as a well-defined, homogeneous, and hyperintense mass on T1-weighted images and hypointense on T2-weighted images [1]. Contrast-enhanced T1-weighted images are the gold standard for imaging of vestibular schwannoma, as they provide the best visualization of the tumor and its relationship to adjacent structures [2]. CT imaging is typically used for preoperative planning and the postoperative phase, as it is not as sensitive as MRI [3]. MRI is also used to monitor tumor growth and response to treatment, as it can detect changes in tumor size and shape [4]. Recent studies have found that the reported sensitivity and specificity of noncontrast MRI is almost equivalent to that of gadolinium-enhanced T1-weighted MRI [5]. Additionally, newer protocols such as FLAIR and DTI may provide additional information and further aid preoperative counseling and surgical planning in the future [6]. Furthermore, deep learning methods have shown state-of-the-art performance for segmenting these structures [7].  Sources:  [1] Vestibular schwannoma. Imaging of acoustic neuroma (vestibular schwannoma). Breakthrough or dilemma?.E Hofmann, L Choné. [2] Vestibular schwannoma. Imaging-based diagnosis of vestibular schwannoma.C Strasilla, V Sychra. [3] Vestibular schwannoma. Current imaging tools for vestibular schwannoma., Linh Dang, Nathan Chin-Yau Tu, Eleanor Y Chan. [4] Vestibular schwannoma. Imaging of the Vestibular Schwannoma: Diagnosis, Monitoring, and Treatment Planning., Steve E J Connor. [5] Vestibular schwannoma. Unsupervised Cross-modality Domain Adaptation for Segmenting Vestibular Schwannoma and Cochlea with Data Augmentation and Model Ensemble, Li, Hao, Hu, Dewei, Zhu, Qibang, Larson, Kathleen E., Zhang, Huahong, Oguz, Ipek. [6] Vestibular schwannoma. EAONO Position Statement on Vestibular Schwannoma: Imaging Assessment. What are the Indications for Performing a Screening MRI Scan for a Potential Vestibular Schwannoma?, Jérôme Waterval, Romain Kania, Thomas Somers. [7] Vestibular schwannoma. Systematic review of the natural history of vestibular schwannoma., Yuhei Yoshimoto. 7. Treatment options for acoustic/vestibular schwannoma include observation, open surgical resection, stereotactic radiosurgery, or fractionated radiotherapy [1]. The indications for surgical treatment of acoustic/vestibular schwannoma include large tumors causing mass effect and obstructive hydrocephalus, tumors that are not amenable to radiosurgery, and tumors that are associated with neurofibromatosis type 2 [2, 3]. Surgery is the preferred treatment for large tumors, as it can achieve the best cytoreductive results and can provide the best facial and hearing nerve outcomes [4]. Radiosurgery is the preferred treatment for smaller tumors, as it can provide strong tumor control and can minimize functional deficit [5]. Fractionated radiotherapy is also an option, with attention to limiting the cochlear radiation dose [4]. The decision to proceed should be individualized and incorporate the multitude of patient- and tumor-specific characteristics known to affect outcome [5]. In the case of neurofibromatosis type 2, the decision should be made in an experienced center [7]. Rehabilitation of auditory, vestibular, motoric, and neurologic functions should also be taken into consideration [7]. Finally, the decision should be based on evidence-based medicine and should respond to the patients individual life situation and preferences [8].  Sources:  [1] Vestibular schwannoma. Acoustic neuroma radiosurgery. Origins, contemporary use and future expectations., D Kondziolka, L D Lunsford, J C Flickinger. [2] Vestibular schwannoma. Vestibular schwannoma management: What is the best option?, S A Rutherford, A T King. [3] Vestibular schwannoma. Acoustic neuroma (vestibular schwannoma) therapy from an oto-rhino-laryngological perspective.K Schwager. [4] Vestibular schwannoma. Contemporary management of acoustic neuromas., Philip V Theodosopoulos, Myles L Pensak. [5] Vestibular schwannoma. Management of Vestibular Schwannoma (Including NF2): Facial Nerve Considerations., Vivian Kaul, Maura K Cosetti. [6] Vestibular schwannoma. Acoustic neurinomes of childhood.A I Krukov, E V Garov, A Yu Ivoilov, E I Zelikovich, A S Kaloshina, V N Zelenkova, S A Gorchakov, A V Zelenkov. [7] Vestibular schwannoma. Vestibular schwannoma. Part 2: therapy, prognosis, and rehabilitation.W Maier, F Hassepaß, A Aschendorff, R Laszig. [8] Vestibular schwannoma. Decision-making of vestibular schwannoma patients., Sabine Müller, Judith Arnolds, Ansel van Oosterhout. 8. The size of acoustic/vestibular schwannoma that is amenable to radiosurgery is typically less than 4 cm3 [3]. Radiosurgery is an effective and safe treatment option for smaller tumors, as demonstrated by a seven-year cohort of patients with VS who were treated at our centre using a linear accelerator-based stereotactic radiosurgery system [1]. In this study, only larger or higher Koos grade VSs were routinely treated with hypofractionated radiosurgery. Another study found that radiosurgery has become an alternative in the treatment of patients with vestibular schwannoma of appropriate size, with high safety level, using low radiation doses, low rate of complications and good tumor control rate in the medium term follow-up [2]. Based on published data on both volume and hearing preservation for both strategies, the authors devised a management recommendation for patients with small vestibular schwannomas, suggesting that early SRS intervention, in contrast to observation, results in long-term tumor control and improved rates of hearing preservation [3].  Sources:  [1] Vestibular schwannoma. Local experience with radiosurgery for vestibular schwannomas and recommendations for management., Ru Xin Wong, Hui Ying Terese Low, Daniel Yat Harn Tan. [2] Vestibular schwannoma. Linear accelerator radiosurgery for the treatment of vestibular schwannoma.R Rodriguez-Mena, J M Gallego-Sanchez, L G Gonzalez-Bonet, F Goig-Revert, C Barcia-Marino, J Rosello-Ferrando. [3] Vestibular schwannoma. The newly diagnosed vestibular schwannoma: radiosurgery, resection, or observation?, Douglas Kondziolka, Seyed H Mousavi, Hideyuki Kano, John C Flickinger, L Dade Lunsford. 9. The surgical approaches used in resecting acoustic/vestibular schwannoma include the translabyrinthine, retrosigmoidal, and subtemporal microsurgical approaches [1]. The translabyrinthine approach is used when the tumor is large and the patient has already experienced some degree of hearing loss. The retrosigmoidal approach is used when the tumor is smaller and hearing preservation is desired [2]. The subtemporal approach is used when the tumor is located in the posterior fossa and the patient has no pre-existing neurologic deficits [3]. All three approaches involve the removal of bone, dural opening, and initial exposure of the tumor [4]. The use of an endoscope has great advantages, as it contributes to the visualization and resection of residual tumor and also reduces the need for cerebellar retraction [5]. The authors present a fully endoscopic resection of a large-sized vestibular schwannoma with facial nerve preservation [6].  Sources:  [1] Vestibular schwannoma. Surgical techniques for treatment of vestibular schwannoma.S Rosahl, D Eßer. [2] Vestibular schwannoma. Acoustic neuroma radiosurgery. Origins, contemporary use and future expectations., D Kondziolka, L D Lunsford, J C Flickinger. [3] Vestibular schwannoma. Retrosigmoid approach to acoustic neuroma (vestibular schwannoma)., R G Ojemann. [4] Vestibular schwannoma. Acoustic neuroma (vestibular schwannoma) therapy from an oto-rhino-laryngological perspective.K Schwager. [5] Vestibular schwannoma. Transcanal surgery for vestibular schwannomas: a pictorial review of radiological findings, surgical anatomy and comparison to the traditional translabyrinthine approach., Matteo Alicandri-Ciufelli, Gaia Federici, Lukas Anschuetz, Giacomo Pavesi, Alessandra Todeschini, Livio Presutti, Daniele Marchioni. [6] Vestibular schwannoma. Endoscopic resection through a retrosigmoid transmeatal approach of a large-sized vestibular schwannoma: operative video and technical nuances., Sebastián J M Giovannini, Guido Caffaratti, Tomas Ries Centeno, Mauro Ruella, Facundo Villamil, Ricardo Marengo, Andrés Cervio. 10. Imaging characteristics and size are important factors in determining the appropriate surgical approach for the excision of vestibular schwannoma. Smaller tumors (<25 mm) are more likely to be managed with observation and serial imaging, stereotactic radiosurgery, or microsurgical resection [1]. Larger tumors (>25 mm) are more likely to require microsurgical resection [2]. Imaging characteristics such as the location of the tumor, its relationship to adjacent structures, and its growth pattern over time can also influence the choice of surgical approach [3, 4]. For example, a study of 28,190 patients with vestibular schwannoma found that the odds ratio for receiving surgery in 2014 was 0.60 compared to those diagnosed in 2004, and the largest increases in observation rates occurred among tumors ≤2 cm [5].  Sources:  [1] Vestibular schwannoma. Imaging of the Vestibular Schwannoma: Diagnosis, Monitoring, and Treatment Planning., Steve E J Connor. [2] Vestibular schwannoma. An update on unilateral sporadic small vestibular schwannoma., Jai Deep Thakur, Anirban Deep Banerjee, Imad Saeed Khan, Ashish Sonig, Cedric D Shorter, Gale L Gardner, Anil Nanda, Bharat Guthikonda. [3] Vestibular schwannoma. Assessment of vestibular schwannoma growth: application of a new measuring protocol to the results of a longitudinal study., E M Stipkovits, K Graamans, G B Vasbinder, J E Van Dijk, F J Beek. [4] Vestibular schwannoma. Imaging-based diagnosis of vestibular schwannoma.C Strasilla, V Sychra. [5] Vestibular schwannoma. Recent Trends in Vestibular Schwannoma Management: An 11-Year Analysis of the National Cancer Database., Solymar Torres Maldonado, James G Naples, Ramie Fathy, Steven J Eliades, John Y K Lee, Jason A Brant, Michael J Ruckenstein. 11. The time course of radiation induced damage to the cochlear nerve following radiosurgery is variable and depends on several factors, including the radiation dose, the volume of the cochlea exposed to radiation, and the patients age and overall health. A review of the literature [1] suggests that radiation-induced hearing loss typically begins to manifest within 6-12 months after radiosurgery, with the most severe hearing loss occurring at 18-24 months [2]. However, some patients may experience hearing loss as early as 3 months after radiosurgery, and the effects of radiation may continue to worsen over time [3].  Sources:  [1] Medulloblastoma, Treatment, Wikipedia. [2] Vestibular schwannoma. Gamma knife radiosurgery for vestibular schwannoma: early hearing outcomes and evaluation of the cochlear dose., John M Lasak, Darren Klish, Thomas C Kryzer, Chris Hearn, John P Gorecki, Grant P Rine. [3] Vestibular schwannoma. Radiation therapy and hearing loss., Niranjan Bhandare, Andrew Jackson, Avraham Eisbruch, Charlie C Pan, John C Flickinger, Patrick Antonelli, William M Mendenhall. 12. Surgery and radiosurgery both carry risks associated with acoustic/vestibular schwannoma. Surgery carries the risk of general anesthesia and surgery of any type, such as myocardial infarction, pneumonia, pulmonary embolism, and infection. Additionally, there is a risk of hydrocephalus, cerebrospinal fluid leak, facial nerve paralysis, facial numbness, hearing loss, ataxia, dysphagia, and major stroke. Radiosurgery carries a risk of cranial nerve morbidity, although this risk has decreased with improvements in dosimetry planning and dose selection [1, 2]. Additionally, there is a risk of hearing loss, which typically begins to manifest within 6-12 months after radiosurgery, with the most severe hearing loss occurring at 18-24 months [3]. Some patients may experience hearing loss as early as 3 months after radiosurgery, and the effects of radiation may continue to worsen over time [3]. Tumor control is higher with surgery than with radiosurgery for small-to-medium size tumors [2], although fractionated radiotherapy has been reported to have a higher rate of hearing preservation than radiosurgery [4].  Sources:  [1] Vestibular schwannoma. Linear accelerator-based radiosurgery for vestibular schwannoma., William A Friedman, Kelly D Foote. [2] Vestibular schwannoma. Audiovestibular symptoms and facial nerve function comparing microsurgery versus SRS for vestibular schwannomas: a systematic review and meta-analysis., Vinod Kumar Yakkala, Marco Mammi, Nayan Lamba, Renuka Kandikatla, Bhaskar Paliwal, Hoda Elshibiny, C Eduardo Corrales, Timothy R Smith, Rania A Mekary. [3] Vestibular schwannoma. A review of treatment modalities for vestibular schwannoma., Benjamin J Arthurs, Robert K Fairbanks, John J Demakas, Wayne T Lamoreaux, Neil A Giddings, Alexander R Mackay, Barton S Cooke, Ameer L Elaimy, Christopher M Lee. [4] Vestibular schwannoma. Acoustic neuroma radiosurgery. Origins, contemporary use and future expectations., D Kondziolka, L D Lunsford, J C Flickinger. 13. Based on the context provided, the treatment options for a patient with a 2cm right acoustic/vestibular schwannoma presenting with right sided tinnitus include observation, open surgical resection, stereotactic radiosurgery, or fractionated radiotherapy [4]. Stereotactic radiosurgery has been proposed as the first line treatment for acoustic tumours and has been particularly advocated in cases of Type 2 Neurofibromatosis (NF2) with bilateral acoustic nerve tumours [1]. However, preserving cochlear nerve function remains a challenging enterprise, and the ideal management that confers the highest chance of hearing preservation remains heavily debated [2]. Given the size of the tumor, surgery may be the best option for preserving hearing, as tumor control is higher with surgery than with radiosurgery for small-to-medium size tumors [3]. Surgery carries risks, including general anesthesia and surgery of any type, cranial nerve morbidity, and hearing loss [3]. Simultaneous cochlear implantation (CI) and microsurgical resection of the tumor has been reported to have positive outcomes due to the binaural benefits, and can be used to treat tinnitus [5].  Sources:  [1] Vestibular schwannoma. Rapid enlargement of a vestibular schwannoma following gamma knife treatment., A W McEvoy, N D Kitchen. [2] Vestibular schwannoma. Retrosigmoid Approach for Vestibular Schwannoma., Mohammed Aref, Katherine Kunigelis, Stephen P Cass, A Samy Youssef. [3] Vestibular schwannoma. Decision-making of vestibular schwannoma patients., Sabine Müller, Judith Arnolds, Ansel van Oosterhout. [4] Vestibular schwannoma. Acoustic neuroma radiosurgery. Origins, contemporary use and future expectations., D Kondziolka, L D Lunsford, J C Flickinger. [5] Vestibular schwannoma. Simultaneous cochlear implantation as a therapeutic option in vestibular schwannoma surgery: case report., Pedro Helo Dos Santos Neto, Johnni Oswaldo Zamponi, Rogério Hamerschmidt, Gislaine Richter Minhoto Wiemes, Marcio S Rassi, Luis A B Borba. 14. There is evidence to suggest that there may be a genetic pre-disposition for acoustic/vestibular schwannoma. In particular, Neurofibromatosis Type 2 (NF2) is a genetic disorder characterized by NF2 mutations that affect the function of a tumor suppressor called merlin, which can lead to bilateral VS and hearing loss [2, 4]. Furthermore, a genome-wide association study identified a risk locus at 9p21.3, which is associated with dysregulation of gene products within the INK4 locus [1, 3]. This suggests that there may be a genetic pre-disposition for acoustic/vestibular schwannoma.  Sources:  [1] Vestibular schwannoma. Double localization of a unilateral sporadic vestibular schwannoma., M Barbara, F Ronchetti, V Manni, S Monini. [2] Vestibular schwannoma. Genome-wide association analysis identifies a susceptibility locus for sporadic vestibular schwannoma at 9p21., Katherine V Sadler, John Bowes, Charlie F Rowlands, Cristina Perez-Becerril, C Mwee van der Meer, Andrew T King, Scott A Rutherford, Omar N Pathmanaban, Charlotte Hammerbeck-Ward, Simon K W Lloyd, Simon R Freeman, Ricky Williams, Cathal John Hannan, Daniel Lewis, Steve Eyre, D Gareth Evans, Miriam J Smith. [3] Vestibular schwannoma. Contemporary Molecular Biology of Sporadic Vestibular Schwannomas: A Systematic Review and Clinical Implications., Hjalte Sass, Per Cayé-Thomasen. [4] Vestibular schwannoma. Genomics, Epigenetics, and Hearing Loss in Neurofibromatosis Type 2., Christine T Dinh, Eric Nisenbaum, Darius Chyou, Carly Misztal, Denise Yan, Rahul Mittal, Juan Young, Mustafa Tekin, Fred Telischi, Cristina Fernandez-Valle, Xue-Zhong Liu. 15. The management of a patient with bilateral acoustic/vestibular schwannoma, who presents with mild-moderate bilateral hearing loss, should be tailored to the individual patient. Generally, the preferred treatment option is surgical resection, as it offers the best chance of preserving hearing. However, due to the risk of facial nerve dysfunction and the potential for multisensory compromise, conservative management may be considered in certain cases, such as those with advanced age or those who refuse surgery [3]. In such cases, regular MRI scans and audiometry should be performed to monitor the tumor growth and hearing function [3]. If the tumor progresses or hearing deteriorates, then surgery should be considered [2]. Hearing preservation attempts should be made during surgery, such as via the middle fossa or retrosigmoid approaches [4], and intraoperative monitoring of hearing should be performed to optimize hearing preservation [4].  Sources:  [1] Vestibular schwannoma. Strategy for the surgical treatment of vestibular schwannomas in patients with neurofibromatosis type 2., Arkadiusz Nowak, Tomasz Dziedzic, Tomasz Czernicki, Przemysław Kunert, Krzysztof Morawski, Kazimierz Niemczyk, Andrzej Marchel. [2] Vestibular schwannoma. Bilateral otoneurological pathology: To operate or not?, Shivun Khosla, Matthew Elliot, Neil Donnelly. [3] Vestibular schwannoma. Conservative management versus surgery for small vestibular schwannomas., Alexis Bozorg Grayeli, Michel Kalamarides, Evelyne Ferrary, Didier Bouccara, Hani El Gharem, Alain Rey, Olivier Sterkers. [4] Vestibular schwannoma. Hearing Preservation in Vestibular Schwannoma Surgery., Joe Saliba, Rick A Friedman, Roberto A Cueva. |
| Expert 1 | 1. It is located between the dorsal plane of the temporal bone, the cerebellum and the pons. 2. Mainly vestibular schwannomas, meningiomas, but also epidermoids, metastasis, endolymphatic sac tumors (especially in Hippel Lindaus), dorsal part of trigeminal schwannomas, ependymomas, medulloblastomas, pilocytic astrocytomas (had a case report), chordomas, chondromas-chondosarcomas, paragangliomas, cholesterol granulomas, hemangioblastoma, ATRTs 3. Mainly adults, middle-aged, seldom children, especially with NF2 schwannomatosis 4. Acute hearing loss, hearing deterioration, vertigo, tinnitus 5. Best with MRI imaging 6. Isodense mass in plain CT, widening of the internal acustic meatus in bone window CT, contrast uptaking mass in contrast CT, T2 hypo/hyperintensiv- T1 isodense-T1 high contrast uptaking mass in MRI 7. Wait and scan, radiosurgery, microsurgical resection, and staged radiotherapy 8. Up to 2-2.5 cm in our hands, we also have a GKN at our institution, if oedema not recommendable 9. Retrosigmoidal, subtemporal 10. Small tumors preferentially treatable with radiosurgery, large ones with resection, if small tumors are very close to the cochlea, surgery may be preferred, because of high hearing preservation with surgery and high chance of loosing hearing because of cochlear radiation dose. Brainstem contact or displacement (Koos III and IV) may cause brainstem oedema 6 months after radiosurgery and may be better treated by resection 11. About 7-10 years until deafness 12. Open surgical resection: wound infection, postop hemorrhage, deafness, facial palsy, brainstem infarction, csf fistulae, deep abscess; Radiosurgery: brainstem oedema, gait disturbances, deafness, facial palsy, tumor progression, malignant transformation 13. Radiosurgery, microsurgical resection, wait and scan 14. NF2 schwannomatosis 15. In small and medium size tumors avastin maybe an option |
| Expert 2 | 1. If the term is used for the space occupied by the cerebellopontine cistern, then it is limited posteromedially by the brainstem, posterolaterally by the petrous portion of the temporal bone and superiorly by the tentorium. While the angle between the superficial transverse fibers of the pons and the fibers of the middle cerebellar peduncle is the anatomical angle. Distinguished anatomical structures are related to this space. Superiorly the trigeminal nerve, inferiorly the lower cranial nerves and in the middle the facial and vestibulocochlear nerves complex with the AICA.  2. Primary neoplasms, most common schwannoma, meningioma, epidermoid, choroid plexus papilloma and exophytic glioma of the brainstem and extensions of ependymomas of the fourth ventricle.   Metastasis and extending tumors of the cranial base, such as chordoma.  Vascular lesions, such as aneurysm, AVM’s, dolichoectatic basilar artery and enlarging cavernous haemangiomas.  Infectious and inflammatory, such as sarcoidosis, tuberculosis and cysticercosis.  3. Adult life, with peak at 65 years-old, with the exception for neurofibromatosis II.  4. According to the size, most with hearing loss, dizziness, and tinnitus.   Large ones may present with neurological dysfunction and hydrocephalus.  5. MRI  6. CT – enlargement of the internal auditory meatus. Heterogeneous enhancement of the mass in the CPA, in large tumors.   MRI – T2 hyperintense lesion in the CPA, possibly cystic, with extension in the internal auditory meatus. Enhancement with gadolinium. For small tumors, FIESTA sequence is helpful.   Medial vestibular schwannoma may not extend laterally into the internal auditory meatus  7. Treatment options include observation, radiosurgery and surgical resection.  Surgery is indicated for:  1) Durable hearing preservation in small tumors; 2) Brainstem decompression in large tumors; and  3) Elimination of progressive growing tumors.  8. I do not prefer radiosurgery regardless of the size.  9. Retrosigmoid, translabyrinthine and middle fossa approaches.   We prefer retrosigmoid approach for the ability to preserve hearing or at least the  cochlear nerve continuity for feature hearing restoration with cochlear implant.  10. The overwhelming majority of tumors of all sizes are well removed through retrosigmoid approach.   Small intracanicular maybe suitable for middle fossa approach.  11. A continuous decline of hearing, over the years to a very low level of preservation at 8 years follow up.  12. CSF leak, infection and hemorrhage are the surgical complications.  13. Surgical resection has a better resolution of tinnitus than radiosurgery. The postoperative cochlear implant may also improve tinnitus outcome after surgical resection for tinnitus.  14. Yes, NF2 gene.  15. This is the most challenging situation and totally individualized life-long management, according to symptoms and tumor progression. |
| Expert 3 | 1. Petrous bone lateral, Pons antero-medial, cerebellum posterior-medial  2. Vestibular schwannomas, meningiomas, epidermoid cysts are the most common  3. Two peaks: between 20-30 (low), between 50-60 (high)  4. Usually vertigo and hearing loss  5. MRI with contrast (gold standard). Also audiogram with speech discrimination and BERA  6. CT isodense, sometimes hypodense due to cystic changes.   MRT T1 with Gd. hyperintense, inhomogeneous. T2 hypointense, sometimes hyperintense due to cystic chances (inhomogeneous)  7. Observation, Radiation (radiosurgery or fractionated), microsurgery  8. Small sizes (T1 – T2)  9. Middle fossa for small tumors with preop. Hearing -> hearing preservation   Translabyrinthine for large tumors with deafness   Retrosigmoid for tumors of all sizes, with or without hearing  10. Middle fossa for small tumors with preop. Hearing -> hearing preservation   Translabyrinthine for large tumors with deafness   Retrosigmoid for tumors of all sizes, with or without hearing  11. 5-10 years  12. Surgery: hearing loss, facial palsy  Radiation: hearing loss, facial spasms, trigeminal symptoms, vertigo  13. It depends on patient’s age. Elderly patients: radiosurgery or microsurgery Young patients: microsurgery is my first choice. Tinnitus improves more likely after surgery than after radiation.  14. Yes. Deletion of NF2 gene on chromosome 22.  15. Observation, or chemotherapy with Bevacizumab, or microsurgery (opening the internal auditory canal and IOM-guided tumor decompression) |
| Expert 4 | 1. I would say angle between cerebellum and pons as name suggests, in terms of anatomical limits: petroclival suture anteriorly, dura posterior to IAC posteriorly (or sigmoid sinus), tentorium/superior petrosal sinus above and jugular foramen below.  2. Main differential – acoustic neuroma, meningioma, epidermoid cyst (actually not a cyst but a tumor); others include trigeminal schwannoma, jugular foramen schwannoma, arachnoid cyst, facial nerve schwannoma  3. Don’t know, would have to look that up. What age groups are you referring to?  4. 1. Hearing loss + tinnitus, brief unsteadiness associated with quick movements, 2. then facial numbness,  3. gait problems are late except when associated with hydrocephalous (communicating with high protein in small tumors), obstructive with large tumors are late.  4. then (late) swallow problems, facial weakness  5. Clinical and radiology  Unilateral sensori neuro hearing loss (acoustic roll over)  MRI fiesta/CISS T2 or gad or both  6. Clinical and radiology  Unilateral sensori neuro hearing loss (acoustic roll over)  MRI fiesta/CISS T2 or gad or both  7. Management options are observation, radiation and surgery  Treatment options are radiation or surgery  Indication for treatment are progressive growth on scans, significant symptom at presentation (large size)/progressive symptoms (excluding hearing loss or tinnitus), I don’t treat for hearing loss – although we try very hard to preserve hearing when we treat tumors, hearing loss not a indication for treatment as most patients have deterioration in hearing after treatment  8. In general 3 cm or less but depends on degree of compression and age of patient. (3 cm may not compress the BS much if atrophy, 2.5 cm tumors may compress significantly and cause facial numbness which radiation would not be as good an option)   Can treat tumors bigger than 3 cm in older patients/frail/comorbidities, those who already have a shunt.  9. Middle fossa – good for hearing preservation but not good for facial nerve (facial nerve in the way), don’t do as often anymore as we don’t treat small tumors and they are observed, mainly tumors that are in the IAC and upto 1cm extension into cp angle/post fossa.   Translab – patients with no hearing, prominent IAC/enlarged IAC component, Don’t like tumors >2.5 cm for translab, not good for big jugular bulb/small mastoid, not good if you can see a pica branch that goes into dura or temporal bone behind IAC on Fiesta scans (they are damaged in translab). – translab is good for short neck/broad shoulders/low tentorium.   Retrosigmoid - Most versatile, potentially hearing preserving, can remove any size tumor. Not as good for short neck/broad shoulders/low tentorium. Also need to look for high jugular bulb.  10. As above, middle fossa is option for mainly IAC tumors, Trans lab for predominantly orlarge IAC component/no large jugular bulb/modest size mastoid/no dural pica branch/better for low tent (and if you can see on scans shoulder and neck anatomy) and not too large (for me), retrosig for any size (don’t like it for low tentorium/short neck)  But other factors non imaging factors are important as outlined in 9.  11. 1-3 years.  12. Risks are different  - Risks of radiation are delayed, does not happen right away.  - 5-10% radiation may not work and tumor keeps growing, requiring more treatment - Radiation has only been in high volume use for 15-20 year and all long term control data is over that time frame, although we think radiation is durable, we don’t have long term data - 1/1000 risk of delayed radiation induced tumors, couple decades after treatment. - Risk to surrounding nerves, (similar to surgery).  • Hearing – 70% ‘preservation’ (for smaller tumors not extending out to the apex of IAC, preservation rate much lower if tumor extends to apex – cochlear dose) but likely 20-30% reduction in function from baseline ( so may not be useful). Hearing loss occurs over 1,2,3 years.  • Facial nerve 1-2 % permanent weakness, can develop weeks or months afterwards. • Facial numbness- if present before tx, some improvement but less likely to improve completely. If not present before treatment, can occur, usually temporary • Unsteadiness – usually improves with tumor stability and other side compensating. • Low likelihood of double vision/swallowing problems, stroke/brain injury • Get to avoid general risk of surgery   Surgery - General risks – anesthetic, bleeding/transfusion/infection/meningitis/pulmonary embolism/dvt/pneumonia/reaction to medicaitons - Specific - Facial nerve – most patients will have some weakness (able to close eye, but slower blink, asymmetric simile, dry eye requiring drops) but will recover over 6 weeks to 6 mos – back to normal or near normal (dependent on tumor size but I usually say 90-95% of the time for tumors 3cm or less). Small proportion will have significant facial weakness – can’t close eye, look bad/crooked face/no movements/like had ‘stroke’/need tape to close eye at night to prevent driness, need eye drops/ointment – things can improve but may take a year or longer for things to start to improve, and will not return back to normal. 1-2 % permanent weakness. This will require help from plastics and ophthalmic surgery for reconstruction, not very good and never looks normal. - Hearing – dependent on hearing level but if class A hearing, I usually say 50% chance of hearing preservation (but what’s preserved is usually 20-30% functionally less – so may not be useful depending on level). All or none in general, unlikely to recover/improve.  - Facial numbness – if present before usually recovers, sometimes doesn’t recover completely. If significant facial numbness before, and doesn’t recover, small risk of deafferentation pain symdrome. Rare to develop numbness after surgery if not present before, usually recovers. IF numbness in face and facial movement (eye closure problem), then significant problem protecting eye…  - Double vision (from 6th nerve or nystagmus) rare – usually gets better, unlikely to require eye balance surgery - Swallow difficulties- rare, feeding tube, unlikely to be permanent  - Stroke /brain stem injury, - rare/unlikely.   Can you die from radiation, probably not, can you die from surgery yes, but probably not much different from being hit by a car when you cross the street, but many people in Canada get hit by cars every day in Canada and if it happens to you it’s 100%  13. Observation  Would repeat scan 6mos to 1 year. If reasonable hearing, suggest hearing aid (fools brain into thinking it can hear and tinnitus may improve – ie tinnitus is a deafferentation problem related to hearing loss  14. yes and no, unlikely if have one acoustic neuroma and no testing further investigations required.  if you have one on both sides, or have a family member who has had brain/acoustic neuromas or have multiple other skin/brain tumors (other schwannomas/menigiomas) you may have Neurofibromatosis and would recommend genetic testing  15. Observation – repeat scan 6 mos to 1 year.  Can make clinical diagnosis of Neurofibromatosis 2, need to tell family members and consider genetic testing for self and children if any. Referral to medical genetics.   Need to do screening scans of spine and rest of brain if MRI was done only to look at IAC.   Need ent involvement if not already to monitor hearing.  Consider support groups  Consider preparing for eventual hearing loss, lip reading courses etc. |
