## Supplementary Table 5 for "neuroGPT-X: Towards an Accountable Expert Opinion Tool for Vestibular Schwannoma"

**Supplementary Table 5.** Response time of expert neurosurgeons and large language models to 15 curated questions. Time is shown as a mean with standard deviation across the 15 questions for each respondent in both seconds and normalized seconds per character.

| **Author** | **Time in Seconds (SD)** | **Normalized Time in Seconds per Character (SD)** |
| --- | --- | --- |
| **GPT Naïve** | 49.04 (10.67) | 0.04 (0.01) |
| **GPT Enriched** | 16.68 (9.29) | 0.02 (0.01) |
| **Gamma** | 63.33 (66.08) | 0.51 (0.14) |
| **Delta** | 30.00 (30.88) | 0.38 (0.82) |
| **Epsilon** | 42.00 (22.1) | 0.69 (0.71) |
| **Zeta** | 134.67 (187.15) | 0.36 (0.24) |
